# DeepPhe-CR: Natural Language Processing Software Services for Cancer Registrar Case Abstraction

**DOI:** 10.1101/2023.05.05.23289524

**Authors:** Harry Hochheiser, Sean Finan, Zhou Yuan, Eric B. Durbin, Jong Cheol Jeong, Isaac Hands, David Rust, Ramakanth Kavuluru, Xiao-Cheng Wu, Jeremy L. Warner, Guergana Savova

**Affiliations:** Department of Biomedical Informatics, University of Pittsburgh, Pittsburgh, PA, USA; Intelligent Systems Program, University of Pittsburgh, Pittsburgh, PA, USA; Boston Childrens’ Hospital, Boston, MA, USA and Harvard Medical School, Boston, MA, USA; Kentucky Cancer Registry, Markey Cancer Center, Lexington, KY, USA; Division of Biomedical Informatics, College of Medicine, University of Kentucky, Lexington, KY, USA; Louisiana Cancer Registry, New Orleans, LA, USA; Lifespan Health System, Providence, RI, USA; Legorreta Cancer Center at Brown University, Providence, RI, USA

**Keywords:** Cancer Informatics, Cancer Registry, Natural Language Processing, Application Programming Interfaces, Data Abstraction

## Abstract

**Objective:** The manual extraction of case details from patient records for cancer surveillance efforts is a resource-intensive task. Natural Language Processing (NLP) techniques have been proposed for automating the identification of key details in clinical notes. Our goal was to develop NLP application programming interfaces (APIs) for integration into cancer registry data abstraction tools in a computer-assisted abstraction setting.

**Methods:** We used cancer registry manual abstraction processes to guide the design of DeepPhe-CR, a web-based NLP service API. The coding of key variables was done through NLP methods validated using established workflows. A container-based implementation including the NLP wasdeveloped. Existing registry data abstraction software was modified to include results from DeepPhe-CR. An initial usability study with data registrars provided early validation of the feasibility of the DeepPhe-CR tools.

**Results:** API calls support submission of single documents and summarization of cases across multiple documents. The container-based implementation uses a REST router to handle requests and support a graph database for storing results. NLP modules extract topography, histology, behavior, laterality, and grade at 0.79-1.00 F1 across common and rare cancer types (breast, prostate, lung, colorectal, ovary and pediatric brain) on data from two cancer registries. Usability study participants were able to use the tool effectively and expressed interest in adopting the tool.

**Discussion:** Our DeepPhe-CR system provides a flexible architecture for building cancer-specific NLP tools directly into registrar workflows in a computer-assisted abstraction setting. Improving user interactions in client tools, may be needed to realize the potential of these approaches. DeepPhe-CR: https://deepphe.github.io/.

## INTRODUCTION

Cancer abstracts based on individual patient records provide the foundation for cancer surveillance efforts at the local, state, and national levels. These reports are generated manually by cancer registrars, who extract descriptors of tumor location, morphology, behavior, and other cancer characteristics from the documentation in the Electronic Health Records (EHRs), e.g. clinical notes, pathology notes and other relevant documentation. Given the complexity and volume of clinical text found in modern EHRs, natural language processing (NLP) techniques present an appealing strategy for optimizing this resource-intensive process. By partially or fully automating the identification of key information, NLP tools have been shown to help registrars identify relevant details,^1–5^ thus potentially speeding the ease and efficiency of data abstraction. To that end, we have adapted our *DeepPhe* (<u>Deep Phe</u>notyping for Oncology Research) system for creating longitudinal patient histories from clinical notes,^6^ to build *DeepPhe-CR* (<u>Deep Phe</u>notyping for <u>C</u>ancer <u>R</u>egistries),^7^ a software-service platform for embedding cancer-optimized NLP into cancer registrar data abstraction tools and workflows in a computer-assisted setting.

Effective cancer surveillance presents a challenge in coordinated data collection. Cancer abstracts often begin at a hospital where a patient is diagnosed or treated for cancer. This initial hospital abstract is collected by skilled registrars, often holding the Certified Tumor Registrar (CTR) certification, according to constantly updated guidelines and data standards published by a variety of organizations such as the National Cancer Institute’s Surveillance Epidemiology and End-Results (SEER) program, the Center for Disease Control’s National Program of Cancer Registries (NPCR), and the American College of Surgeons’ Commission on Cancer (CoC).

Contributions from all these standards agencies are coordinated by the North American Association of Central Cancer Registries (NAACCR) into an annual data standards release, describing the procedures for extracting key details such as tumor site, morphology, stage, first-course of treatment, and survival^8^. Once collected, hospital abstracts are submitted to the state central cancer registry. At the state cancer registry, hospital abstracts and other clinical documents are consolidated into a single patient-level document (although abstracting may be further complicated for patients with multiple primary cancers) using established workflows supported by registry software tools such as SEER*DMS^9^, and are then submitted to national programs such as SEER and NPCR. With an increasingly fluid data ecosystem, cancer registrars are expected to synthesize information from a variety of clinical notes, pathology reports, and other sources of narrative text for submission to many different local and national agencies, according to increasingly complex data collection standards.

As identifying and coding relevant content in clinical notes can be a highly laborious process, partially or fully automated approaches based on NLP techniques have been implemented as a means of increasing efficiency and accuracy (as reviewed by Savova, et al. ^10^ and Wang et al.^11^). A variety of NLP methods have been applied to a number of challenges in extracting cancer information from clinical text, including treatments^12^, recurrences^13^, and other attributes.^14^ Recent efforts have explored the application of these techniques to SEER registry tasks, including the application of multi-task convolutional neural networks (CNNs) to a large set of electronic documents from the Louisiana Tumor Registry,^1^ the use of CNNs to extract information from pediatric cancer pathology notes,^2^ mapping of concepts in pathology notes to ICD-O-3 codes,^4^ and the use of transfer learning techniques to apply models trained in one registry to notes from other registries.^3^

Since 2014, we have been developing the DeepPhe system to extract and visualize longitudinal patient histories from clinical text to combine with the structured data from electronic medical record (EMRs). Based on the Apache Clinical Text and Knowledge Extraction System (cTAKES) NLP platform,^15^ DeepPhe combines focused NLP for identifying and coding key cancer variables with capabilities for summarization to provide views at multiple granularities, from individual mentions to high-level summaries.^6^ Extracted results can be displayed and interpreted at both the cohort and individual patient and cancer levels using our web-based visual analytics tool.^16^

A broad range of prior efforts have used NLP techniques to extract phenotypes from the EHR clinical narrative.^10^ Perhaps most similar to DeepPhe, recent efforts by Alawad, et al.^1^ and Yoon et al.^5^ have successfully applied multi-task deep convolutional learning to extract attributes including site, laterality, behavior, histology, and grade. However, these tools work in batch modes, processing large corpora of textual documents and assembling outputs for a subsequent analysis. These methods also have relied on a “bronze” standard for ground truth where the predictions are modeled using the values found in abstracted registry reports associated with the collection of pathology documents for a tumor, rather than gold standard codes derived from each pathology document itself. This approach is not without limitations: performance is good for more common sites but suffers for rarer sites.

As pointed out above, DeepPhe-CR (**<u>DeepPhe</u>** for **<u>C</u>**ancer **<u>R</u>**egistries) builds on DeepPhe but was designed to support registrar data abstraction processes, which rely upon manual review of patient’s EMR documents to identify key cancer details, which are then entered into appropriate forms in web-based interfaces. Tools that close the gap between these workflow steps add NLP outputs to registry workflows, potentially bringing improvements in task completion times, accuracy, and user satisfaction. Specifically, NLP-enhanced abstraction tools might extract relevant concepts, pre-populating appropriate fields in registry interfaces, thus providing registrars with faster access to relevant values and guiding their review of the notes. Although this approach – known as computer-assisted clinical coding, has been explored in a number of fields,^17^ including recent applications to cancer registry abstraction,^1–4^ reports to date have focused on information extraction and classification tasks for full automation, with little attention paid to the details of how these techniques might be integrated into registry tools and workflows in a computer-assisted setting.

To further explore the possible benefits of integrating NLP into cancer registry workflows, our team of informaticians, clinicians, epidemiologists, and registrars collaborated to understand registrar needs, built the modified version of DeepPhe -- DeepPhe-CR -- providing REST^18^ API calls designed to support integration with registry software in a computer-assisted setting. Here, we describe the goals and design of the system, and lessons learned through the interactions with registrars at multiple SEER registries.

## METHODS

### Basic Requirements

Initial design discussions with cancer registrars identified several criteria that must be met for a successful integration of DeepPhe-CR with registry software. As the widely used SEER*DMS software and other prominent registry tools are web-based, integration through client-side HTTP calls was determined to be the most straightforward approach. Specifically, we decided to expose DeepPhe-CR functionality via the REST software architectural style API^18^, capable of submitting documents and retrieving results, including the type and location of relevant text spans (indicated by start and end character indices).

We also identified the importance of providing this REST API as a single point of access. The principal DeepPhe software stack (which we modified for DeepPhe-CR) involves multiple components, including a processing pipeline and a database for result storage. To simplify integration while providing maximal flexibility for future evolution, the design of DeepPhe-CR aims to hide these components behind a single-entry point. To simplify adoption, the DeepPhe-CR software is provided as a set of Docker containers designed for easy installation and configuration.

Finally, because our goal is computer-assisted abstraction we identified several steps necessary to support registrars in their review and coding of clinical documents. The DeepPhe-CR output include spans and their represented classes for registry-pertinent extracted variables. Registrars review documents as presented in text windows in a graphical user interface. A “pre-annotation” setup for using NLP results to facilitate abstraction is needed to support this effort. This configuration activates the NLP modules to identify text spans associated with extracted items and color-codes each span of the relevant data item type in the text window.

Discussions with registrars also led to the identification of primary site (topography, major and minor), histology, behavior, laterality, grade, and stage as the key variables to be extracted from the notes (Table 1). In addition to the required elements, DeepPhe-CR extracts certain biomarkers, which are items of growing interest to the registry community. To balance registrars’ needs for highly-accurate extraction against the challenges of building highly effective clinical NLP tools, we established an initial goal of extraction of each of these attributes at F1 scores of 0.75 or better to demonstrate initial feasibility for computer-assisted abstraction. This goal of 0.75 F1 is guidance provided by the Cancer Registries based on their previous work on efficiency within a computer-assisted setting. F1 is the harmonic mean of precision/positive predictive value and recall/sensitivity and is the classic metric for reporting overall NLP system performance.

**Table 1:** Cancer data elements to be extracted; AJCC: American Joint Committee on Cancer; NAACCR: North American Association of Central Cancer Registries; TNM: tumor, node, metastasis.

| NAACCR Data item | NAACCR Item Number |
| --- | --- |
| Primary Site (topography – major and minor) | 400 |
| Histology | 522 |
| Behavior Code | 523 |
| Laterality | 410 |
| Grade | 440 |
| AJCC Clinical TNM Stage | 1001, 1002, 1003, 1004 |
| AJCC Pathological TNM Stage | 1011, 1012, 1013, 1014 |

### Inquiries, API Development, and Prototype

A series of discussions and software demonstrations served to introduce the DeepPhe-CR software development team to registrar workflow practices. Insights from these discussions were used to develop the draft specification for the REST API. Based on this draft, we developed an initial version of the DeepPhe-CR software stack. The specification document and the DeepPhe-CR tools were iteratively refined until the necessary functionality was complete to allow integration into registry software. Discussions with two independent registry software developer teams validated this approach.

### Information Extraction

A serially executed NLP workflow was used to develop the approaches for extracting the required data items. Two domain experts and an informatician (GS) developed detailed annotation guidelines and piloted them on a set of 30 patients. Disagreements were tracked and discussed. As a result, the annotation guidelines were adjusted to address the areas of disagreement. Inter-annotator agreement was computated at 0.77-1 kappa on an additional set of 30 patients. Gold standard annotations for 1560 randomly selected patients with several common cancer types – breast, lung, colorectal, ovarian, and prostate as well as a rare type of cancer (pediatric brain cancer) were created based on the finalized annotation guidelines and split into training, development, and test sets. The data were provided by two SEER cancer registries (Kentucky and Louisiana). Pediatric brain cancer was included as it represents a rare type cancer thus data sparcity. The original DeepPhe ontology was modified to the ICD-O to create the DeepPhe-CR ontology. Thus, the DeepPhe-CR consists of the existing DeepPhe approaches^6^ with the DeepPhe-CR ontology as the backbone. The train split was used to identify missing concepts from the DeepPhe-CR ontology and the development split – to validate the ontology extensions. DeepPhe-CR was evaluated on the held-out test split. Table 2 provides the distribution across types of cancers and train/development/test splits.

**Table 2:** Data distribution (number of patients) across common types of cancers (breast, lung, prostate, colorectal, ovarian) and a rare type of cancer (pediatric brain cancer)

|  | **Breast** | **Lung** | **Prostate** | **Colorectal** | **Ovarian** | **Pediatric Brain** | **Total** |
| --- | --- | --- | --- | --- | --- | --- | --- |
| **Train** | 231 | 198 | 178 | 90 | 90 | 120 | 907 |
| **Development** | 72 | 55 | 48 | 30 | 29 | 41 | 234 |
| **Test** | 134 | 120 | 106 | 30 | 29 | 39 | 419 |
| **Total** | 437 | 373 | 332 | 150 | 148 | 200 | 1560 |

### Integration Example, User Study, and Generalization

Upon completion of the initial API, implementations, and methods development, the tools were integrated into the Kentucky Cancer Registry’s Cancer Patient Data Management System (CPDMS), a robust and stable software platform developed and implemented across the state.^19^ This initial implementation was designed to demonstrate the feasibility of including DeepPhe-CR in a registry-scale software system in a computer-assisted abstraction setting. In a series of recorded sessions, three members of the project team who were skilled with cancer data abstraction were provided a brief introduction to the augmented CPDMS tool. They then used both the original CPDMS tool and the enhanced version to abstract the information for cancer cases, each supported by documents of varying length and relevant context. Accuracy, task completion time, and qualitative observations were used to descriptively assess the utility of the revised workflow.

Discussions with registry staff from two additional states (Massachusetts and Louisiana) were used to verify generalizability of the workflow to new contexts. Similarly, new gold standard datasets were created to account for data from the Louisiana Tumor Registry, which was used to further evaluate the system.

## RESULTS

### Inquiries, API Development, and Prototype

Our discussions of functional requirements and our observations of registrar practices confirmed our early design decisions, including the provision of REST calls through a single point of access, and the need for including spans and associated classes within the responses. Our inquiries into the registry workflows revealed that registry processes are driven by individual documents, not patients. Each document is reviewed as it comes in, at which time it is either associated with an existing patient/tumor or a new patient/tumor record is created. Thus, it is desirable for the DeepPhe-CR API to include calls for submitting individual documents and receiving appropriate results. Finally, security was identified as a priority. Although registry software will generally be hosted in a secure environment, some form of authorization should be associated with each REST call.

To address these needs, as well as the requirements for a single-point of access and ease of installation, we developed a Docker-based architecture involving two containers: a reverse proxy container and a container for the core document processing. The reverse proxy container provides the single point of access through a web server, handing calls off to the document processing container. The use of this proxy server adds minimal overhead while providing the flexibility needed for further evolution, allowing the addition of new calls, or even the reallocation of the document processing services into multiple containers, without requiring any changes to client systems. Security is provided through an HTTP Bearer-Token that must be provided to authenticate each request.

The core document processing system is a modified version of the original DeepPhe system, enhanced to support the single-document approach needed by registry workflows. Although not immediately required by the registries, support for patient-level document submission (as offered by the original DeepPhe) was included as well. Together, these features are implemented through several key components found in the document processing container: 1) The core NLP functionality; 2) a Neo4j graph database server augmented with a DeepPhe-CR extension for storing and retrieving results from individual documents, as necessary for patient summarization; 3) a summarizer module; and 4) a query processor capable of handling requests for summarization and other stored data. An overview of the DeepPhe-CR architecture is given in Figure 1.

**Figure 1:**
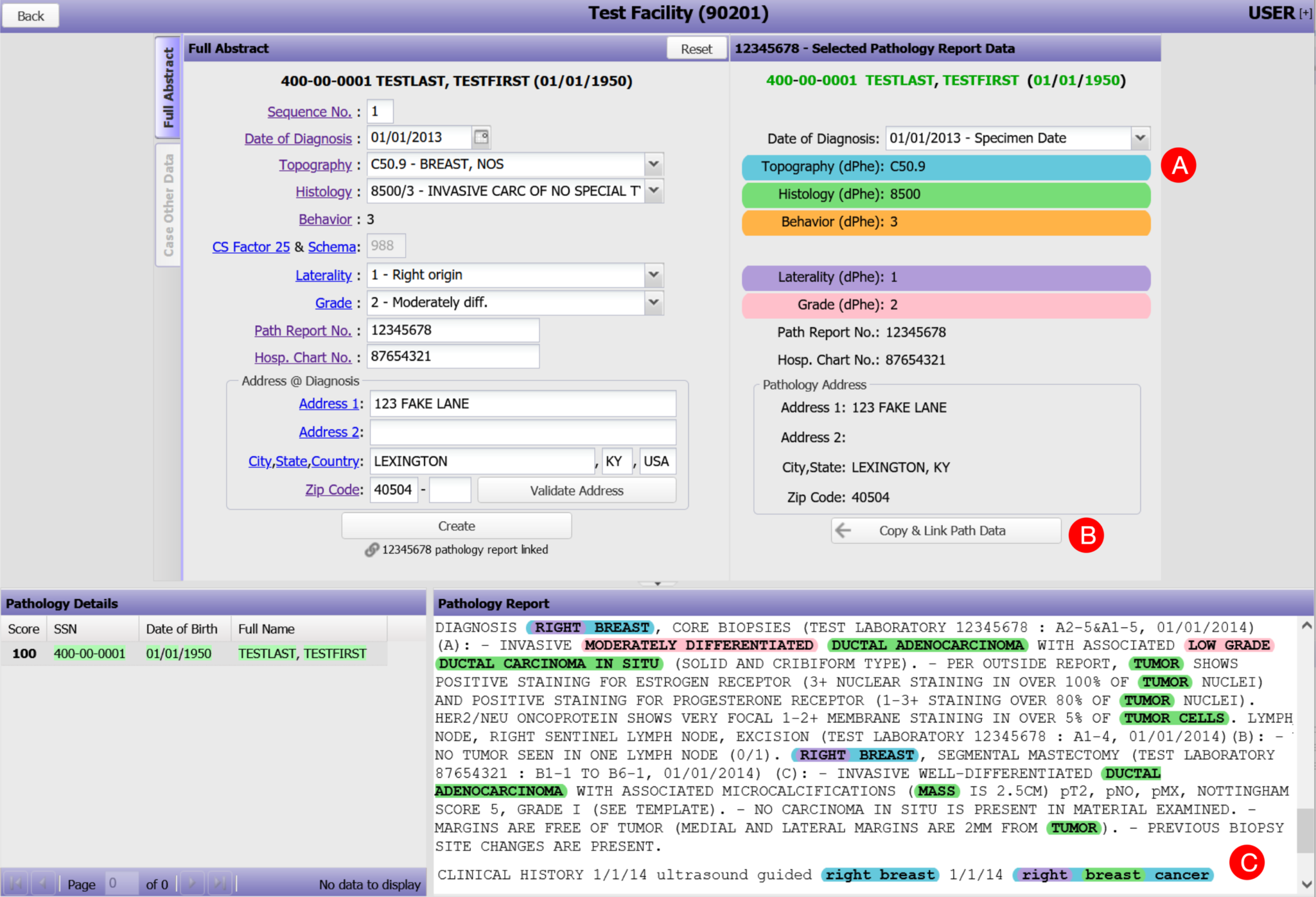
The data abstraction screen from the Cancer Patient Data Management System, augmented to support suggested data items as extracted from clinical text by DeepPhe-CR: (A) document-level topography, histology, behavior, laterality, and grade values are shown color-coded and labelled with appropriate ICD-O/NAACCR codes; (B) Suggested items and demographics can be copied to the in-progress abstract with a single click; (C) The clinical text is highlighted with color codes indicating text spans associated with the five summary values displayed in the suggestion area above (A). Thus, “Right breast cancer” is associated with topography C50.9, “breast” with histology 8500, and “right” with laterality 1.

The DeepPhe-CR API supports four calls (Table 3). The simplest approach, designed to meet the most important need identified by the registries, takes a single document and returns the NLP-extracted data. Alternatively, one or more documents can be submitted and processed, with results stored for summarization and retrieval via a subsequent API call. This submission can take one of two forms – immediate return of results, and queueing for future processing. In the case of a single document, the summary will contain only the data found in the document, and this workflow is essentially equivalent to single document submission and summary retrieval. If multiple documents for the same patient are submitted, a requested patient summary will address items contained in all documents. Sample calls and returned results are given in Supplemental Appendix A..

**Table 3:** API calls for submitting information to the DeepPhe-CR services and retrieving results

| API Call | Description | Return value |
| --- | --- | --- |
| summarizeOneDoc | Summarize a single document, no caching | JSON containing single-document neoplasm summary |
| summarizePatientDoc | Summarize a single document, cache the document for patient summary to be retrieved on subsequent call | JSON containing single-document neoplasm summary results |
| queuePatientDoc | Cache the document for patient summary to be retrieved on subsequent call. Used to submit multiple documents prior for summarization via summarizePatients call. | JSON containing a Boolean value indicating that the the document has been submitted. |
| summarizePatient | Retrieve summarized patient information | JSON containing all results extracted for a patient, including a cross-document summary |

DeepPhe-CR’s Docker-based containerized implementation is straightforward to install, requiring only a few commands. DeepPhe-CR source code (all open source), installation instructions, documentation API calls and tools for managing the Docker containers, including execution of integration tests, can be found on the DeepPhe-CR release GitHub site, reachable through: https://deepphe.github.io.

### Information Extraction

The information extracted by the DeepPhe-CR is listed in Table 1. As an extension of the original DeepPhe system, DeepPhe-CR specifically addresses the extraction of values for these required variables from clinical free text of varied types (e.g. clinical notes, radiology reports, pathology notes, etc.). As was described above, the DeepPhe-CR approaches are the same as DeepPhe’s^6^ – an end-to-end hybrid pipeline combining symbolic and machine learners driven by knowledge represented in its DeepPhe-CR backbone ontology. The machine learners are for the tasks of sentence splitting and part-of-speech tagging which we take from Apache cTAKES; therefore we did not retrain them. DeepPhe-CR results on the held-out test split are in Table 4. The results demonstrate that DeepPhe-CR achieves high F1 scores on both the common (colorectal, prostate, breast, ovary and lung) and rare (pediatric brain) types of cancers. Note that because DeepPhe-CR assigns a value for each required category of topography, histology, behavior, laterality and grade, the F1 equates to accuracy thus precision and recall values are the same (assuming no missing values in the gold annotations).

**Figure 1:**
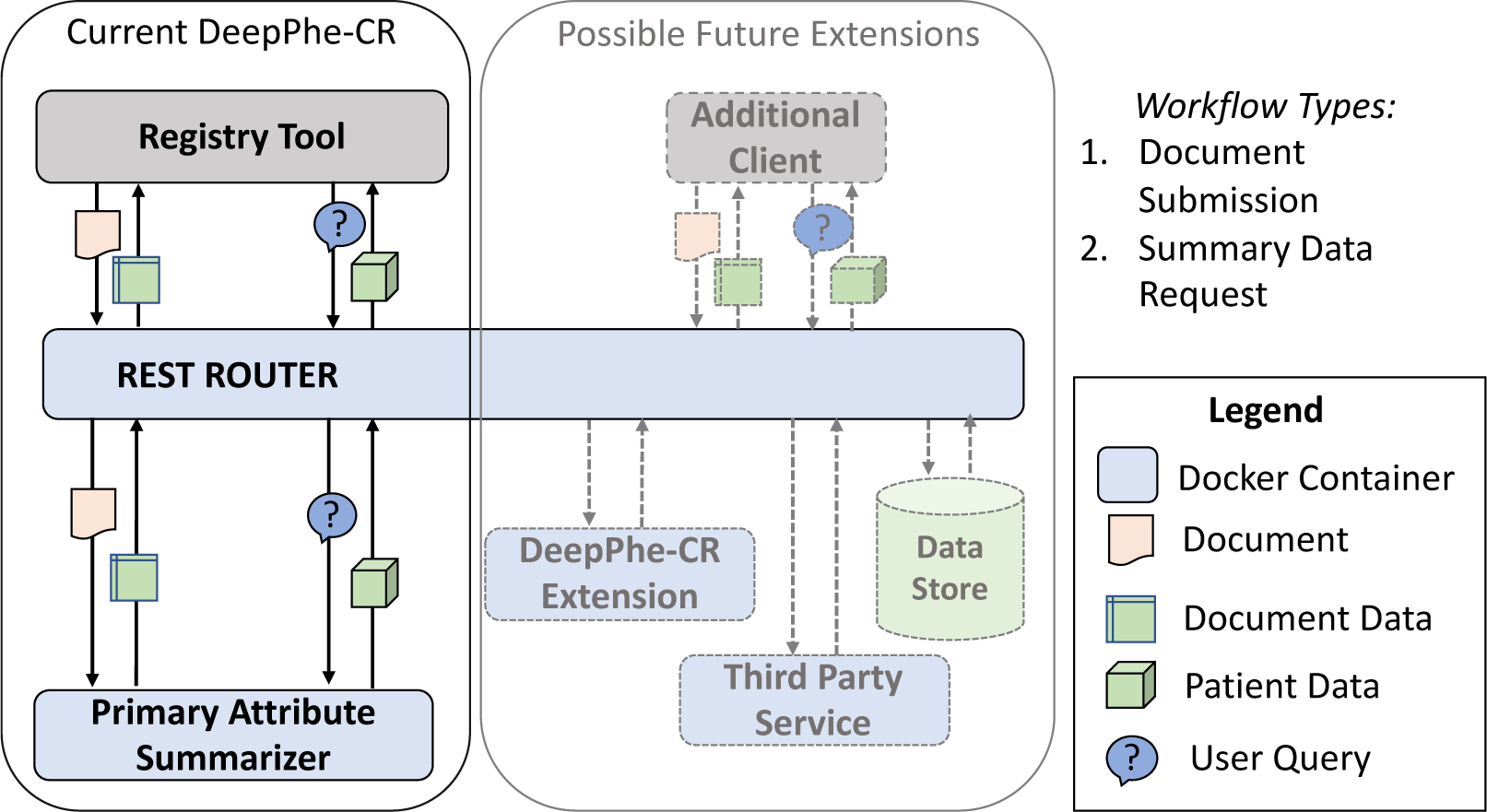
The DeepPhe-CR architecture

**Table 4:**
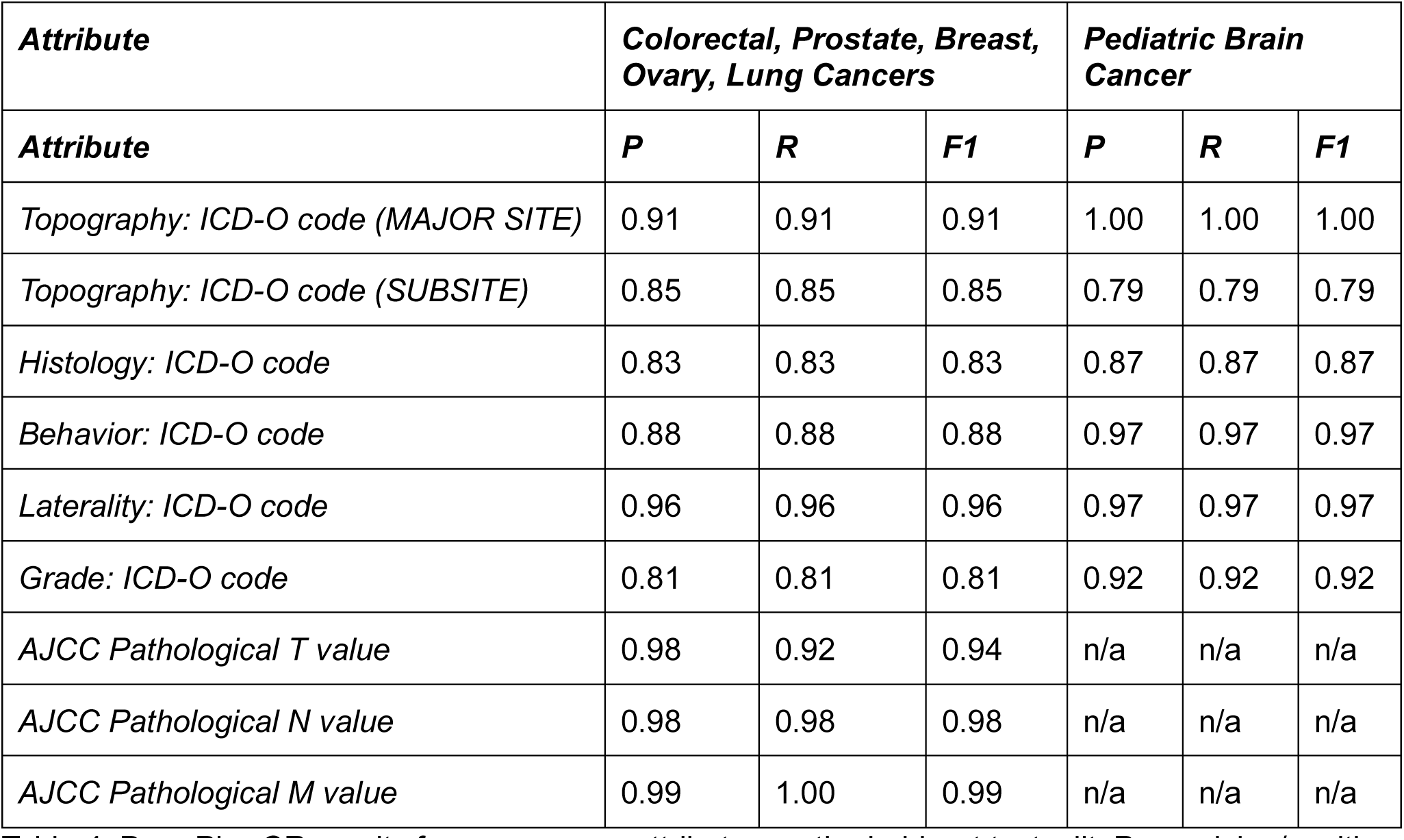
DeepPhe-CR results for cancer core attributes on the held-out testsplit. P=precision/positive predictive value; R=recall/sensitivity; F1=harmonic mean of precision and recall

The DeepPhe-CR ontology includes representation of biomarkers and their relations to other classes, such as cancer types. This allows the system to extract and summarize biomarkers of interest and associate them with other extracted information. Biomarker classes are mostly assembled from NCIt,^20^ though some protein classes come from Human Phenotype Ontology (HPO).^21^ Custom relations were introduced between appropriate cancer branches and biomarkers, e.g. breast cancer and ER, PR, HER2. The system can identify relations of almost 150 types. DeepPhe-CR achieved 0.90 F1 for biomarker extraction with precision/positive predictive value 0.91 and recall/sensitivity 0.89 on the test split from Table 2.

### Integration Example, User Study, and Generalization

The integration of the DeepPhe-CR features into the CPDMS tool involved the addition of two new features to the basic CPDMS data capture screen. A new set of controls on the right-hand side of the screen displays suggested items as extracted by DeepPhe-CR. At the bottom of the screen, a text box displays the document from which information is extracted, with spans highlighted in colors matching those used to label the suggested items. Selected items can be copied to the data annotation inputs with a single click (Figure 2).

In an initial usability study, two participating registrars experienced with cancer data abstraction used the enhanced CPDMS/DeepPhe-CR tool to annotate cancer documents. Although participants differed in how quickly they were able to use the tools, both were able to use the tool appropriately and expressed enthusiasm for the enhancements.

The relationship between task completion time with and without DeepPhe-CR annotations was mixed. For one of the two participants, task completion time was significantly faster with the DeepPhe-CR annotations (with DeepPhe-CR average time 31.7 sec, standard deviation 14.0 sec; without DeepPhe-CR average time 59.7 seconds, standard deviation 14.9 sec, Wilcoxon’s W, p < 0.05). No significant difference was found for the other participant. This may be due to the need to review the document to avoid omitting any relevant information: even when they felt confident about the results extracted by DeepPhe-CR, participants carefully reviewed documents in search of any additional information that may have been missed by the NLP tools.

## DISCUSSION

Cancer surveillance is built on the foundation of cancer registry data abstraction work, involving manually reading and extracting information from clinical text. This careful interpretation of clinical free-text is a resource-intensive process, requiring significant person-power from trained experts who identify and code key items of interest. NLP tools capable of automating or semi-automating the identification of these key details have the potential to improve registrar efficiency and accuracy. Improving the efficiency of registry workflows also facilitates the expansion of the registry dataset to also include additional information, such as genomic reports, without additional staff resources.

Our DeepPhe-CR system provides a flexible architecture for building cancer-specific NLP tools directly into registrar workflows in a computer-assisted abstraction setting. Based on a familiar containerized implementation and REST-like^18^ architectures, these tools can easily be installed and accessed by web-based registry tools, thus minimizing changes to familiar workflows and encouraging adoption. An initial deployment within the Kentucky Cancer Registry’s CPDMS provides a demonstration of the feasibility of this approach.

Our aspirational goal is to develop methods to enable the near-complete automation of many cancer registry data abstraction tasks. Although the performance of the DeepPhe-CR NLP components meets the set goal of 0.75 F1 for a computer-assisted abstraction, the goal for full automation is in the 95-98% F1 range, as informally set forward by SEER and others. For a subset of the documents, that high level F1 is already achieved. Methods for identifying these documents with high confidence need to be developed thus introducing complete automation for a portion of the incoming data. Our goal is to facilitate cancer registrar efforts, providing greater user satisfaction and confidence in resulting abstraction in a human-computer interaction mode. Thus, as of now, DeepPhe-CR tools provide useful input that helps registrars complete their work within the computer-assisted abstraction setting. We also note that DeepPhe-CR is a modular architecture with inherent pluggable utility to registry software and can be enhanced with more accurate NLP methods.

The use of DeepPhe-CR annotations as a tool for supporting manual abstracting and coding is consistent with our observations during our user study of the integration of DeepPhe-CR into the Kentucky CPDMS. Although participants were enthusiastic about the NLP assistance and showed clear signs of using the extracted attributes, we did not see any consistent indication that the NLP tools helped to reduce the time required to complete abstraction tasks. Some of this effect was clearly due to the need to verify system feedback --users were establishing their trust in the system -- as participants regularly scrolled through documents to find highlighted text corresponding to DeepPhe-CR suggestions. System suggestions that were either omitted (i.e., when DeepPhe-CR was unable to identify an attribute of a tumor) or incorrect caused problems, as users had to read the note to find appropriate text. We believe that continuous and prolonged use of DeepPhe-CR will aid in user gains in familiarity and trust of the system and of the relatively high performance of the methods for selected attributes.

Enhancements to the registry software user interface and workflow to provide additional support for the integration of NLP output predictions might provide additional gains. The integration of confidence scores displayed to the registrars for each DeepPhe-CR extracted variable was the most requested feature to add. Such scores might help focus registrar attention on those suggested items that have the greatest uncertainty and are currently under development. User interface enhancements, such as showing the context in which extracted items were found, might help minimize the effort required to verify NLP suggestions. The design of the DeepPhe-CR API also has the potential to provide additional gains, if supported through appropriate revisions to registry workflows. Specifically, DeepPhe-CR’s ability to summarize multiple pathology reports might reduce the effort needed to manually link details across reports. However, revisions to data abstraction workflows and tools will be necessary to realize these improvements.

More advanced functionality might engage registrars in providing feedback that might be used to improve the NLP tools. Such tools might allow users to indicate when an extracted attribute was correct and to suggest alternate values for incorrect results, providing feedback that might be incorporated into dynamically evolving methods for information extraction. Further work might be needed to determine appropriate approaches for evaluating the impact of the NLP-augmented abstraction process. Although reductions in the time required for document abstraction might appear to be the most obvious potential benefit, other metrics such as subjective satisfaction and accuracy should be considered. Even if subsequent studies continue to suggest that the DeepPhe-CR annotations do not reduce task completion time, improvements in these alternative measures might be sufficient evidence to justify adoption of the NLP-aided approach to cancer data abstraction.

Limitations of this work include the relatively small sample size in terms of the number of participants in the usability study. Although the document corpus is adequate for methods development, it remains possible that results on unseen datasets will not reach the F1 scores seen in the testing set. The validation of the NLP approaches with clinical data from additional registries along with additional types of cancers and of the tools with users from those sites will be needed to demonstrate broader generalizable utility.

## CONCLUSION

The use of NLP to assist in the extraction of required reporting of cancer attributes and streamlining cancer data abstraction processes is an appealing possibility. Our DeepPhe-CR system provides a containerized set of abstraction tools supported by a REST API, providing infrastructure suitable for integration into registry software. Accurate methods and a use case demonstrate the feasibility of this approach, while also demonstrating the need for improved methods. The DeepPhe-CR tools are available at https://deepphe.github.io.

## Data Availability

The software described in this paper is available online at https://deepphe.github.io

## ACKNOWLEDGMENTS

David Harris and Lisa Witt provided vital annotation help and assistance in user-testing prototype tools. Brent Mumphrey, Susan Gershman, and Hung Tran provided useful feedback to the development of the tools described in this paper.

## Supplemental Appendix: Examples of DeepPhe-CR calls and results

### 1. Introduction

We use manually created synthetic notes (available at https://github.com/DeepPhe/dphe-examples/reports) to demonstrate the use of DeepPhe-CR API calls to process clinical notes.

DeepPhe-CR uses a simple RESTful architecture, following common conventions similar to many other APIs.

### 2. Synthetic Sample Report

Progress note: fake_patient4_doc1_NOTE.txt

Report ID…………………1,doc1

Patient ID pt567567567

Patient Name………………Fake Patient4

Principal Date…………….20110310 1025

Record Type NOTE

Patient DOB 01/15/1949

Reason for Visit: Patient is a 62 year old post-menopausal female with a long history of lumpy breasts. She felt a mass on her right breast on self-examination that she hadn’t noticed before. She also felt another lump in her left breast which she described as hard and painful.

Interim History: The patient is a 62 year old female who presents for evaluation of the masses. The patient’s recent mammography on March 1, 2011 revealed a lobulated mass in the upper inner quadrant of the right breast at 2 o’clock position that measured 1.2×3.0 cm. Mammography also revealed a nodular mass in the upper inner quadrant of the left breast at 11 o’clock position that measured 2 cm in diameter.

Ultrasound evaluation was performed on the area of the right breast and a 1.1×2.2×3.3 cm hypoechoic mass is seen in the 2 o’clock region. Ultrasound of the left breast at 11 o’clock revealed a 1.1×1.2×1.2 cm mass. Ultrasound of the right axillary lymph nodes was performed showing multiple thickened lymph nodes. Ultrasound of the left axilla revealed normal appearing lymph nodes.

Past History: She has a history of left ovarian carcinoma, treated with total abdominal hysterectomy, bilateral salpingo-oophorectomy in July 2002. She received neoadjuvant chemotherapy with Taxol and Carboplatin.

Physical Exam: Health appearing female in NAD. HEENT: NC/AT, PERRL, EOMI, Sclera non icteric NECK: Supple, no masses

LUNGS: CTA, no wheezes, rales, rhonchi CARDIAC: RRR. No murmurs, rubs or gallops ADB: Soft, NT, ND, No HSM

EXT: No clubbing, cyanosis or edema

Impression: A 62 year old female with bilateral breast masses seen on mammography and ultrasound scheduled for core biopsy of the right breast mass as well as the left breast nodule. She will return to discuss further treatment options once pathology is finalized.

### 3. Structure of API calls

All API calls follow a common structure, including:

· An “Authorization: Bearer” header field with the authorization token.

· A “Content-Type: text/plain” header field

· A unique identifier for the document and/or patient, as appropriate

· Document text sent as a binary payload

All of the API calls conform to a common basic format:

PUT/GET http://[hostanme]:[port]/DeepPhe/[API call name]/doc/[doc name] [document text]

where [hostname]:port specify the server name and port - localhost:8080 by default.

### 4. Example call: summarizeOneDoc – single document summarization

To summarize a single document, the only necessary identifier is the document ID. No patient ID is needed. Following the pattern From section 3, the pattern is as follows:

PUT http://localhost:8080/DeepPhe/summarizeOneDoc/doc/doc1 [document text]

Using the command-line cURL tool (https://curl.se/) and the note stored in a file named “fake_patient4_doc1_NOTE.txt”, we can make this call as follows:

curl -i -X GET http://localhost:8080/deepphe/summarizeDoc/doc/doc1 \

-H “Content-Type: text/plain” \

-H “Authorization: Bearer AbCdEf123456” \

--data-binary “@fake_patient4_doc1_NOTE.txt”

Note that this call uses the default Bearer token installed/provided with the system.

### 5. DeepPhe-CR output

Results are returned as structured JSON blocks containing lists and nested name, value dictionaries. A complete example is given in Section 7.

The summarizeOneDoc call given in Section 4, when applied to the note give in Section 2, will be used to describe the output.

Differences in output formats for specific calls are described alongside those calls. The top-level output of a single document Id contains the following items:

id the document Id submitted to the call

patient A description of the patient and key information extracted from the document

neoplasms a list of summarized tumors.

These subsections are described in Sections 5.1 and 5.2 The full JSON output is given in Section 7.

#### 5.1 Patient

The patient field contains:

id defaults to the docid for the summarizeOneDoc call

type document -type “Clinical Note”

date date when processed

text the text of the note

sections a list of sections in the document. Each section has an ID, a type, and a start and end character offset

mentions a list of individual concepts mentioned in the document

relations a list of relations between the mentions

corefs a list of coreferences, currently unused

##### 5.1.1 Mentions

A mention is a word or other string of characters identified by DeepPhe-CR as being relevant to one of the classes of information to be extracted. Each mention has several attributes:

id: a unique identifier, generated by the system

classUri: a unique label corresponding to the DeepPhe-CR concept associated with the text

noteid: an internal identifier for the note

begin: the starting character offset for the word/string

end: the ending character offset for the word/string

There are also several binary values that can modify each mention: negated, uncertain, generic, conditional, historic. These fields will have values of true as applicable. Thus, if the mention was associated with the phrase “patient has a history of cancer”, the historic flag would be set to True.

An example for a mention of a breast mass:

{

“id”: “doc1_M_3”,

“classUri”: “Mass”, “noteId”: “doc1_884540155”, “begin”: 428,

“end”: 432, “negated”: false, “uncertain”: false,

“generic”: false, “conditional”: false, “historic”: false, “temporality”: “”

}

##### 5.1.2 Relations

A relation is an indication of a semantically meaningful link between two mentions. Relationships are given in as (subject,predicate,target) triples, indicating that the subject is in a relationship of type predicate with the target

type: A descriptor of the predicate of the relationship. Examples include hasDiagnosis, hasTumorExtent, relationships for clinical stage indicators, etc.

sourceId: the subject of the relationship, referring to an ID of one of the mentions in the document.

targetId: the ID of the mention of the object of the relationship. Thus, for example the relationship:

{

“type”: “hasTumorExtent”, “sourceId”: “doc1_M_3”, “targetId”: “doc1_M_1”

}

indicates that mention doc1_M_3 is a mention of a tumor that has an extent given by mention

doc1_M_1.

#### 5.2 Neoplasms

The neoplasms record is a list of the identified cancers. Each cancer is represented by the following items:

id: a unique, system-generated identifier for the neoplasm

classUri: a label for the class of neoplasm involved – for example, “Mass_In_Breast”

attributes: a list of attributes of the neoplasm

##### 5.2.1 Neoplasm Attributes

Neoplasm attributes provide summarized descriptions of each cancer. Each attribute contains several metadata items:

id: a unique, system-generated identifier for the attribute

classUri: a label for the class of attribute involved – for example, “Mass_In_Breast”

name: a name for the attribute – for example, “behavior” or “histology”

value: the extracted value for the attribute

confidenceFeatures: a list of integers used internally by the DeepPhe system to

calculate confidence values. These features are opaque and not designed to be used otherwise.

Each attribute also contains three lists of *evidence:* directEvidence, indirectEvidence, and notEvidence, corresponding respectively to those items that clearly support a summary, those that provide less direct support, and those that are not evidence for that item. Each evidence item is a copy of a specific mention (see A.5.1.1) from the text.

### 6. Complete API Specification

Note: As described in Section 3, all calls require “Content-Type: text/plain” and “Authorization: Bearer ” headers.

#### 6.1 summarizeOneDoc

This basic call simply returns the result of summarizing the provided document.

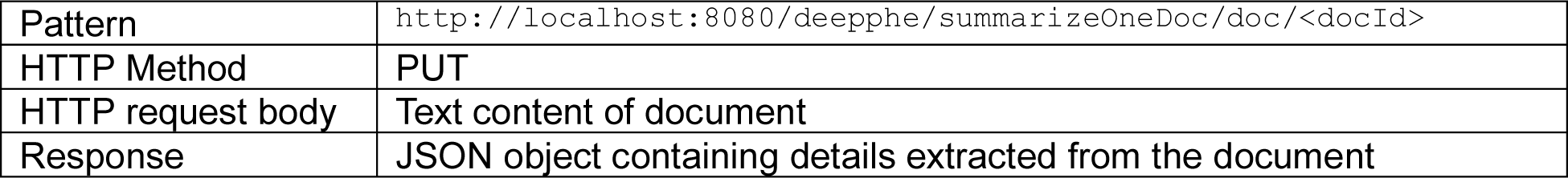

#### 6.2 summarizePatientDoc

This call will process a document associated with a given patient, retaining extracted information for a subsequent summary. The information returned will include the neoplasm summary for the given document, without any of the sections, mentions, or relations.

Summary information for the patient can be extracted by a subsequent call to summarizePatient, using the specified. See Section 6.4 for details.

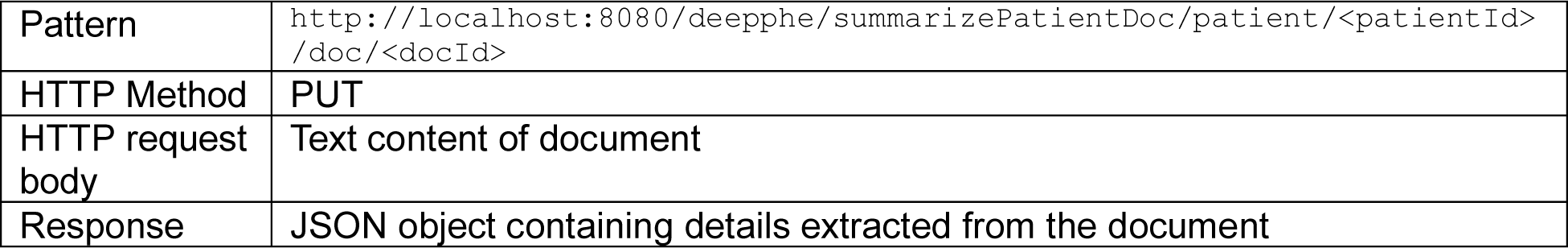

#### 6.3 queuePatientDoc

This call is similar to summarizePatientDoc, but the summary is not directly returned. The only response is a JSON block indicating that the document has been queued.

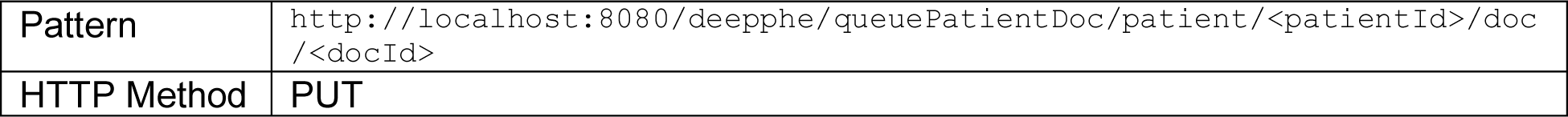
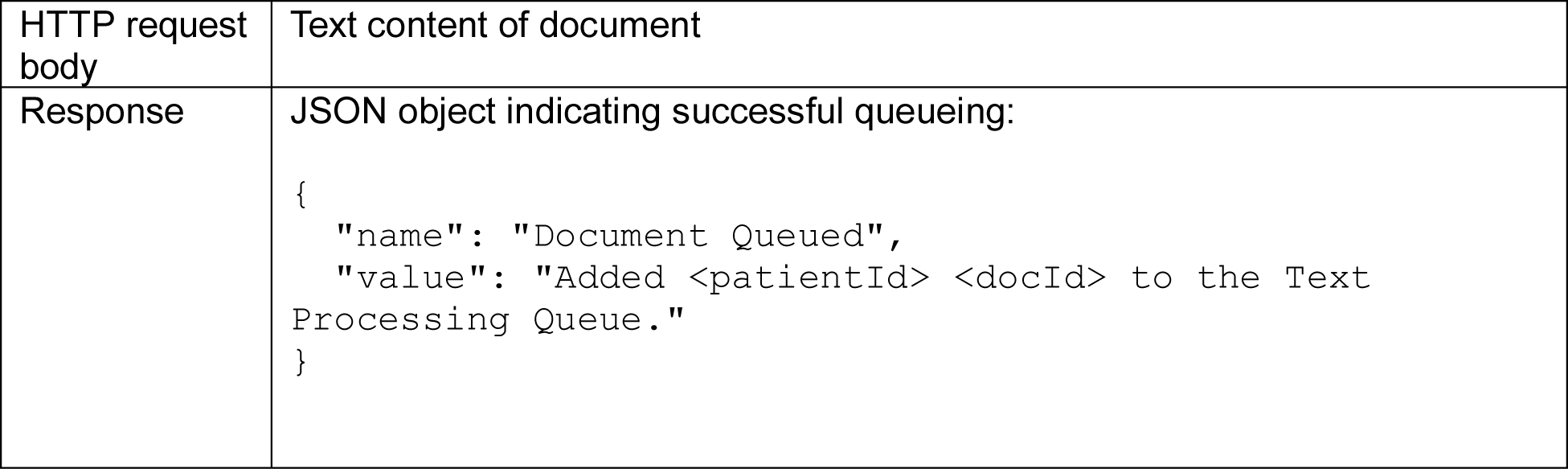

#### 6.4 summarizePatient

SummarizePatient builds a summary of all information submitted for a given patient, through prior calls to summarizePatientDoc and queuePatientDoc. If only one document has been submitted for the patient, the results are equivalent to what would be returned for a single call (for that document) to summarizeDoc.

Note that this call uses GET, unlike the other calls, which use PUT to submit document text.

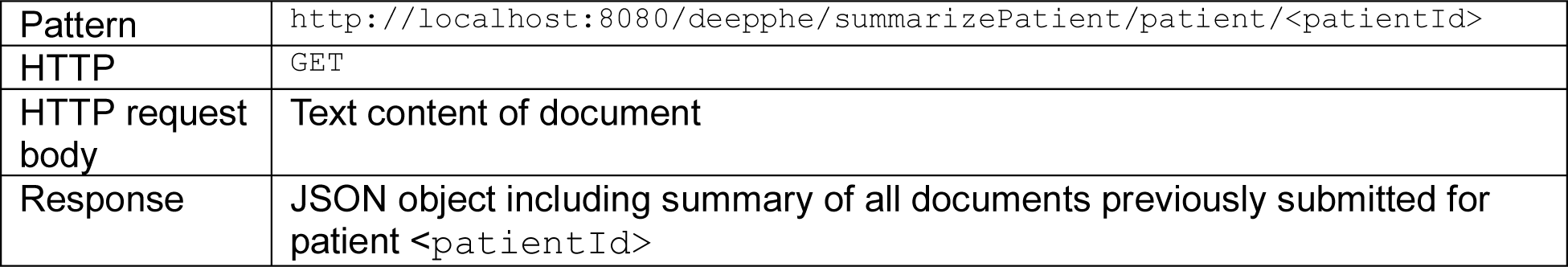

## 7. Complete Example

The following JSON is output for the summarizeOneDoc call executed against the example file given in section 2.1.

{

“id”: “doc1”, “patient”: {

“id”: “doc1”,

“name”: “doc1”, “notes”: [

{

“id”: “doc1_884540155”,

“name”: “doc1”,

“type”: “Clinical Note”, “date”: “202305030238”,

“episode”: “unknown”, “text”:

“\u003d\u003d\u003d\u003d\u003d\u003d\u003d\u003d\u003d\u003d\u003d\u003d\u003d\u003d\ u003d\u003d\u003d\u003d\u003d\u003d\u003d\u003d\u003d\u003d\u003d\u003d\u003d\u003d\u0 03d\u003d\u003d\u003d\u003d\u003d\u003d\u003d\u003d\u003d\u003d\u003d\u003d\u003d\u003 d\u003d\u003d\u003d\u003d\u003d\u003d\u003d\u003d\u003d\u003d\u003d\u003d\u003d\u003d\ u003d\u003d\u003d\u003d\u003d\u003d\u003d\u003d\u003d\u003d\nReport ID…………………1,doc1\nPatient ID pt567567567\nPatient

Name ………………Fake Patient4\nPrincipal Date 20110310 1025\nRecord Type NOTE\nPatient DOB……………….01/15/1949\n\nReason for Visit: Patient is a 62 year old post-menopausal female with a long history of lumpy breasts. She felt a mass on her right breast on self-examination that she hadn’t noticed before. She also felt another lump in her left breast which she described as hard and painful.\n\nInterim History: The patient is a 62 year old female who presents for evaluation of the masses. The patient’s recent mammography on March 1, 2011 revealed a lobulated mass in the upper inner quadrant of the right breast at 2 o’clock position that measured 1.2×3.0 cm.

Mammography also revealed a nodular mass in the upper inner quadrant of the left breast at 11 o’clock position that measured 2 cm in diameter. \nUltrasound evaluation was performed on the area of the right breast and a 1.1×2.2×3.3 cm hypoechoic mass is seen in the 2 o’clock region. Ultrasound of the left breast at 11 o’clock revealed a 1.1×1.2×1.2 cm mass. Ultrasound of the right axillary lymph nodes was performed showing multiple thickened lymph nodes. Ultrasound of the left axilla revealed normal appearing lymph nodes.\n\nPast History: She has a history of left ovarian carcinoma, treated with total abdominal hysterectomy, bilateral salpingo-oophorectomy in July 2002. She received neoadjuvant chemotherapy with Taxol and Carboplatin.\n\nPhysical Exam: Health appearing female in NAD. \nHEENT: NC/AT, PERRL, EOMI, Sclera non icteric\nNECK: Supple, no masses\nLUNGS: CTA, no wheezes, rales, rhonchi\nCARDIAC: RRR. No murmurs, rubs or gallops\nADB: Soft, NT, ND, No HSM\nEXT: No clubbing, cyanosis or edema\n\n\nImpression: A 62 year old female with bilateral breast masses seen on mammography and ultrasound scheduled for core biopsy of the right breast mass as well as the left breast nodule. She will return to discuss further treatment options once pathology is finalized.\n\n”,

“sections”: [

{

“id”: “doc1_S_1”,

“type”: “Pittsburgh Header”,

“begin”: 67,

“end”: 594

},

{

“id”: “doc1_S_2”,

“type”: “Interim History”,

“begin”: 611,

“end”: 1608

},

{

“id”: “doc1_S_3”,

“type”: “Examination”,

“begin”: 1623,

“end”: 1871

},

{

“id”: “doc1_S_4”,

“type”: “Impression”,

“begin”: 1883,

“end”: 2137

}

],

“mentions”: [

{

“id”: “doc1_884540155_M_1”,

“classUri”: “Lymph_Node”,

“confidence”: 0.0, “noteId”: “doc1_884540155”,

“begin”: 1308,

“historic”: false, “temporality”: “”

},

{

“id”: “doc1_884540155_M_2”,

“classUri”: “Mass”,

“confidence”: 0.0, “noteId”: “doc1_884540155”,

“begin”: 1215,

“end”: 1219,

“negated”: false,

“uncertain”: false,

“generic”: false,

“conditional”: false,

“historic”: false,

“temporality”: “”

},

{

“id”: “doc1_884540155_M_3”,

“classUri”: “Mass_In_Breast”,

“confidence”: 0.0,

“noteId”: “doc1_884540155”,

“begin”: 402,

“end”: 415,

“negated”: false,

“uncertain”: true,

“generic”: false,

“conditional”: false,

“historic”: true,

“temporality”: “”

},

{

“id”: “doc1_884540155_M_5”,

“classUri”: “Mass_In_Breast”,

“confidence”: 0.0,

“noteId”: “doc1_884540155”,

“begin”: 2007,

“end”: 2018,

“negated”: false,

“uncertain”: false,

“generic”: false,

“conditional”: false,

“historic”: false,

“temporality”: “”

},

{

“id”: “doc1_884540155_M_4”,

“classUri”: “Heart”,

“confidence”: 0.0,

“noteId”: “doc1_884540155”,

“begin”: 1765,

“end”: 1772,

“negated”: false,

“uncertain”: false,

“generic”: false,

“conditional”: false,

“historic”: false,

“temporality”: “”

},

{

“id”: “doc1_884540155_M_6”,

“classUri”: “Axillary_Lymph_Node”,

“noteId”: “doc1_884540155”,

“begin”: 1246,

“end”: 1266,

“negated”: false,

“uncertain”: false,

“generic”: false,

“conditional”: false,

“historic”: false,

“temporality”: “”

},

{

“id”: “doc1_884540155_M_7”,

“classUri”: “Left_Axilla_Proper”,

“confidence”: 0.0,

“noteId”: “doc1_884540155”,

“begin”: 1340,

“end”: 1351,

“negated”: false,

“uncertain”: false,

“generic”: false,

“conditional”: false,

“historic”: false,

“temporality”: “”

},

{

“id”: “doc1_884540155_M_8”,

“classUri”: “Right”,

“confidence”: 0.0,

“noteId”: “doc1_884540155”,

“begin”: 440,

“end”: 445,

“negated”: false,

“uncertain”: false,

“generic”: false,

“conditional”: false,

“historic”: false,

“temporality”: “”

},

{

“id”: “doc1_884540155_M_9”,

“classUri”: “Upper_inner_Quadrant”,

“confidence”: 0.0,

“noteId”: “doc1_884540155”,

“begin”: 911,

“end”: 931,

“negated”: false,

“uncertain”: false,

“generic”: false,

“conditional”: false,

“historic”: false,

“temporality”: “”

},

{

“id”: “doc1_884540155_M_10”,

“classUri”: “Breast”,

“confidence”: 0.0,

“noteId”: “doc1_884540155”,

“begin”: 2007,

“historic”: false,

“temporality”: “”

},

{

“id”: “doc1_884540155_M_11”,

“classUri”: “Right”,

“confidence”: 0.0,

“noteId”: “doc1_884540155”,

“begin”: 1062,

“end”: 1067,

“negated”: false,

“uncertain”: false,

“generic”: false,

“conditional”: false,

“historic”: false,

“temporality”: “”

},

{

“id”: “doc1_884540155_M_12”,

“classUri”: “Right”,

“confidence”: 0.0,

“noteId”: “doc1_884540155”,

“begin”: 1240,

“end”: 1245,

“negated”: false,

“uncertain”: false,

“generic”: false,

“conditional”: false,

“historic”: false,

“temporality”: “”

},

{

“id”: “doc1_884540155_M_13”,

“classUri”: “Breast”,

“confidence”: 0.0,

“noteId”: “doc1_884540155”,

“begin”: 807,

“end”: 813,

“negated”: false,

“uncertain”: false,

“generic”: false,

“conditional”: false,

“historic”: false,

“temporality”: “”

},

{

“id”: “doc1_884540155_M_14”,

“classUri”: “Bilateral”,

“confidence”: 0.0,

“noteId”: “doc1_884540155”,

“begin”: 1909,

“end”: 1918,

“negated”: false,

“uncertain”: false,

“generic”: false,

“conditional”: false,

“historic”: false,

“temporality”: “”

},

{

“id”: “doc1_884540155_M_15”,

“classUri”: “Right”,

“noteId”: “doc1_884540155”,

“begin”: 801,

“end”: 806,

“negated”: false,

“uncertain”: false,

“generic”: false,

“conditional”: false,

“historic”: false,

“temporality”: “”

},

{

“id”: “doc1_884540155_M_16”,

“classUri”: “Neck”,

“confidence”: 0.0,

“noteId”: “doc1_884540155”,

“begin”: 1702,

“end”: 1706,

“negated”: false,

“uncertain”: false,

“generic”: false,

“conditional”: false,

“historic”: false,

“temporality”: “”

},

{

“id”: “doc1_884540155_M_17”,

“classUri”: “Left”,

“confidence”: 0.0,

“noteId”: “doc1_884540155”,

“begin”: 1427,

“end”: 1431,

“negated”: false,

“uncertain”: true,

“generic”: false,

“conditional”: false,

“historic”: true,

“temporality”: “”

},

{

“id”: “doc1_884540155_M_18”,

“classUri”: “Epithelial_Ovarian_Cancer”,

“confidence”: 0.0,

“noteId”: “doc1_884540155”,

“begin”: 1432,

“end”: 1449,

“negated”: false,

“uncertain”: false,

“generic”: false,

“conditional”: false,

“historic”: true,

“temporality”: “”

},

{

“id”: “doc1_884540155_M_19”,

“classUri”: “Lung”,

“confidence”: 0.0,

“noteId”: “doc1_884540155”,

“begin”: 1726,

“end”: 1731,

“historic”: false,

“temporality”: “”

},

{

“id”: “doc1_884540155_M_20”,

“classUri”: “Dysplastic_Nevus”,

“confidence”: 0.0,

“noteId”: “doc1_884540155”,

“begin”: 1650,

“end”: 1653,

“negated”: false,

“uncertain”: false,

“generic”: false,

“conditional”: false,

“historic”: false,

“temporality”: “”

},

{

“id”: “doc1_884540155_M_21”,

“classUri”: “Mass”,

“confidence”: 0.0,

“noteId”: “doc1_884540155”,

“begin”: 428,

“end”: 432,

“negated”: false,

“uncertain”: false,

“generic”: false,

“conditional”: false,

“historic”: false,

“temporality”: “”

},

{

“id”: “doc1_884540155_M_22”,

“classUri”: “Mass”,

“confidence”: 0.0,

“noteId”: “doc1_884540155”,

“begin”: 528,

“end”: 532,

“negated”: false,

“uncertain”: false,

“generic”: false,

“conditional”: false,

“historic”: false,

“temporality”: “”

},

{

“id”: “doc1_884540155_M_24”,

“classUri”: “Breast”,

“confidence”: 0.0,

“noteId”: “doc1_884540155”,

“begin”: 1168,

“end”: 1174,

“negated”: false,

“uncertain”: false,

“generic”: false,

“conditional”: false,

“historic”: false,

“temporality”: “”

},

{

“id”: “doc1_884540155_M_23”,

“classUri”: “Lobular_Mass”,

“noteId”: “doc1_884540155”,

“begin”: 751,

“end”: 765,

“negated”: false,

“uncertain”: false,

“generic”: false,

“conditional”: false,

“historic”: false,

“temporality”: “”

},

{

“id”: “doc1_884540155_M_25”,

“classUri”: “Position_Of_Phenotypic_Abnormality”,

“confidence”: 0.0,

“noteId”: “doc1_884540155”,

“begin”: 827,

“end”: 835,

“negated”: false,

“uncertain”: false,

“generic”: false,

“conditional”: false,

“historic”: false,

“temporality”: “”

},

{

“id”: “doc1_884540155_M_26”,

“classUri”: “Breast”,

“confidence”: 0.0,

“noteId”: “doc1_884540155”,

“begin”: 408,

“end”: 415,

“negated”: false,

“uncertain”: false,

“generic”: false,

“conditional”: false,

“historic”: true,

“temporality”: “”

},

{

“id”: “doc1_884540155_M_27”,

“classUri”: “Breast”,

“confidence”: 0.0,

“noteId”: “doc1_884540155”,

“begin”: 1919,

“end”: 1925,

“negated”: false,

“uncertain”: false,

“generic”: false,

“conditional”: false,

“historic”: false,

“temporality”: “”

},

{

“id”: “doc1_884540155_M_28”,

“classUri”: “Upper_inner_Quadrant”,

“confidence”: 0.0,

“noteId”: “doc1_884540155”,

“begin”: 773,

“historic”: false,

“temporality”: “”

},

{

“id”: “doc1_884540155_M_30”,

“classUri”: “Left”,

“confidence”: 0.0,

“noteId”: “doc1_884540155”,

“begin”: 1340,

“end”: 1344,

“negated”: false,

“uncertain”: false,

“generic”: false,

“conditional”: false,

“historic”: false,

“temporality”: “”

},

{

“id”: “doc1_884540155_M_29”,

“classUri”: “Breast”,

“confidence”: 0.0,

“noteId”: “doc1_884540155”,

“begin”: 2039,

“end”: 2045,

“negated”: false,

“uncertain”: false,

“generic”: false,

“conditional”: false,

“historic”: false,

“temporality”: “”

},

{

“id”: “doc1_884540155_M_31”,

“classUri”: “Left”,

“confidence”: 0.0,

“noteId”: “doc1_884540155”,

“begin”: 939,

“end”: 943,

“negated”: false,

“uncertain”: false,

“generic”: false,

“conditional”: false,

“historic”: false,

“temporality”: “”

},

{

“id”: “doc1_884540155_M_34”,

“classUri”: “Left”,

“confidence”: 0.0,

“noteId”: “doc1_884540155”,

“begin”: 1163,

“end”: 1167,

“negated”: false,

“uncertain”: false,

“generic”: false,

“conditional”: false,

“historic”: false,

“temporality”: “”

},

{

“id”: “doc1_884540155_M_33”,

“classUri”: “Sclera”,

“noteId”: “doc1_884540155”,

“begin”: 1683,

“end”: 1689,

“negated”: false,

“uncertain”: false,

“generic”: false,

“conditional”: false,

“historic”: false,

“temporality”: “”

},

{

“id”: “doc1_884540155_M_32”,

“classUri”: “Position_Of_Phenotypic_Abnormality”,

“confidence”: 0.0,

“noteId”: “doc1_884540155”,

“begin”: 965,

“end”: 973,

“negated”: false,

“uncertain”: false,

“generic”: false,

“conditional”: false,

“historic”: false,

“temporality”: “”

},

{

“id”: “doc1_884540155_M_35”,

“classUri”: “Mass_In_Breast”,

“confidence”: 0.0,

“noteId”: “doc1_884540155”,

“begin”: 2039,

“end”: 2052,

“negated”: false,

“uncertain”: false,

“generic”: false,

“conditional”: false,

“historic”: false,

“temporality”: “”

},

{

“id”: “doc1_884540155_M_37”,

“classUri”: “Left”,

“confidence”: 0.0,

“noteId”: “doc1_884540155”,

“begin”: 540,

“end”: 544,

“negated”: false,

“uncertain”: false,

“generic”: false,

“conditional”: false,

“historic”: false,

“temporality”: “”

},

{

“id”: “doc1_884540155_M_36”,

“classUri”: “Mass”,

“confidence”: 0.0,

“noteId”: “doc1_884540155”,

“begin”: 899,

“historic”: false,

“temporality”: “”

},

{

“id”: “doc1_884540155_M_38”,

“classUri”: “Mass”,

“confidence”: 0.0,

“noteId”: “doc1_884540155”,

“begin”: 1107,

“end”: 1111,

“negated”: false,

“uncertain”: false,

“generic”: false,

“conditional”: false,

“historic”: false,

“temporality”: “”

},

{

“id”: “doc1_884540155_M_41”,

“classUri”: “Bilateral”,

“confidence”: 0.0,

“noteId”: “doc1_884540155”,

“begin”: 1494,

“end”: 1503,

“negated”: false,

“uncertain”: false,

“generic”: false,

“conditional”: false,

“historic”: false,

“temporality”: “”

},

{

“id”: “doc1_884540155_M_40”,

“classUri”: “Ovary”,

“confidence”: 0.0,

“noteId”: “doc1_884540155”,

“begin”: 1432,

“end”: 1439,

“negated”: false,

“uncertain”: false,

“generic”: false,

“conditional”: false,

“historic”: true,

“temporality”: “”

},

{

“id”: “doc1_884540155_M_39”,

“classUri”: “Lymph_Node”,

“confidence”: 0.0,

“noteId”: “doc1_884540155”,

“begin”: 1378,

“end”: 1389,

“negated”: false,

“uncertain”: false,

“generic”: false,

“conditional”: false,

“historic”: false,

“temporality”: “”

},

{

“id”: “doc1_884540155_M_42”,

“classUri”: “Left”,

“noteId”: “doc1_884540155”,

“begin”: 2034,

“end”: 2038,

“negated”: false,

“uncertain”: false,

“generic”: false,

“conditional”: false,

“historic”: false,

“temporality”: “”

},

{

“id”: “doc1_884540155_M_44”,

“classUri”: “Right”,

“confidence”: 0.0,

“noteId”: “doc1_884540155”,

“begin”: 2001,

“end”: 2006,

“negated”: false,

“uncertain”: false,

“generic”: false,

“conditional”: false,

“historic”: false,

“temporality”: “”

},

{

“id”: “doc1_884540155_M_43”,

“classUri”: “Breast”,

“confidence”: 0.0,

“noteId”: “doc1_884540155”,

“begin”: 1068,

“end”: 1074,

“negated”: false,

“uncertain”: false,

“generic”: false,

“conditional”: false,

“historic”: false,

“temporality”: “”

},

{

“id”: “doc1_884540155_M_45”,

“classUri”: “Breast”,

“confidence”: 0.0,

“noteId”: “doc1_884540155”,

“begin”: 944,

“end”: 950,

“negated”: false,

“uncertain”: false,

“generic”: false,

“conditional”: false,

“historic”: false,

“temporality”: “”

},

{

“id”: “doc1_884540155_M_46”,

“classUri”: “Breast”,

“confidence”: 0.0,

“noteId”: “doc1_884540155”,

“begin”: 545,

“historic”: false,

“temporality”: “”

},

{

“id”: “doc1_884540155_M_47”,

“classUri”: “Breast”,

“confidence”: 0.0,

“noteId”: “doc1_884540155”,

“begin”: 446,

“end”: 452,

“negated”: false,

“uncertain”: false,

“generic”: false,

“conditional”: false,

“historic”: false,

“temporality”: “”

}

],

“relations”: [

{

“type”: “Disease_Has_Associated_Anatomic_Site”,

“sourceId”: “doc1_884540155_M_18”,

“targetId”: “doc1_884540155_M_4”,

“confidence”: 40.25

},

{

“type”: “Disease_Has_Associated_Anatomic_Site”,

“sourceId”: “doc1_884540155_M_18”,

“targetId”: “doc1_884540155_M_7”,

“confidence”: 55.70000076293945

},

{

“type”: “Disease_Has_Associated_Anatomic_Site”,

“sourceId”: “doc1_884540155_M_18”,

“targetId”: “doc1_884540155_M_9”,

“confidence”: 46.79999923706055

},

{

“type”: “Disease_Has_Associated_Anatomic_Site”,

“sourceId”: “doc1_884540155_M_18”,

“targetId”: “doc1_884540155_M_10”,

“confidence”: 21.25

},

{

“type”: “Disease_Has_Associated_Anatomic_Site”,

“sourceId”: “doc1_884540155_M_18”,

“targetId”: “doc1_884540155_M_13”,

“confidence”: 44.400001525878906

},

{

“type”: “Disease_Has_Associated_Anatomic_Site”,

“sourceId”: “doc1_884540155_M_18”,

“targetId”: “doc1_884540155_M_16”,

“confidence”: 35.25

},

{

“type”: “Disease_Has_Associated_Anatomic_Site”,

“sourceId”: “doc1_884540155_M_18”,

“targetId”: “doc1_884540155_M_19”,

“confidence”: 43.25

},

{

“type”: “Disease_Has_Associated_Anatomic_Site”,

“sourceId”: “doc1_884540155_M_18”,

“targetId”: “doc1_884540155_M_24”,

“confidence”: 52.20000076293945

},

{

“type”: “Disease_Has_Associated_Anatomic_Site”,

“sourceId”: “doc1_884540155_M_18”,

“targetId”: “doc1_884540155_M_26”,

“confidence”: 11.5

},

{

“type”: “Disease_Has_Associated_Anatomic_Site”,

“sourceId”: “doc1_884540155_M_18”,

“targetId”: “doc1_884540155_M_27”,

“confidence”: 24.75

},

{

“type”: “Disease_Has_Associated_Anatomic_Site”,

“sourceId”: “doc1_884540155_M_18”,

“targetId”: “doc1_884540155_M_28”,

“confidence”: 44.0

},

{

“type”: “Disease_Has_Associated_Anatomic_Site”,

“sourceId”: “doc1_884540155_M_18”,

“targetId”: “doc1_884540155_M_29”,

“confidence”: 19.5

},

{

“type”: “Disease_Has_Associated_Anatomic_Site”,

“sourceId”: “doc1_884540155_M_18”,

“targetId”: “doc1_884540155_M_33”,

“confidence”: 46.5

},

{

“type”: “Disease_Has_Associated_Anatomic_Site”,

“sourceId”: “doc1_884540155_M_18”,

“targetId”: “doc1_884540155_M_40”,

“confidence”: 100.0

},

{

“type”: “Disease_Has_Associated_Anatomic_Site”,

“sourceId”: “doc1_884540155_M_18”,

“targetId”: “doc1_884540155_M_43”,

“confidence”: 49.70000076293945

},

{

“type”: “Disease_Has_Associated_Anatomic_Site”,

“sourceId”: “doc1_884540155_M_18”,

“targetId”: “doc1_884540155_M_45”,

“confidence”: 47.20000076293945

},

{

“type”: “Disease_Has_Associated_Anatomic_Site”,

“sourceId”: “doc1_884540155_M_18”,

“targetId”: “doc1_884540155_M_46”,

“confidence”: 11.5

},

{

“type”: “Disease_Has_Associated_Anatomic_Site”,

“sourceId”: “doc1_884540155_M_18”,

“targetId”: “doc1_884540155_M_47”,

“confidence”: 11.5

},

{

“type”: “Disease_Has_Primary_Anatomic_Site”,

“sourceId”: “doc1_884540155_M_18”,

“targetId”: “doc1_884540155_M_40”,

“confidence”: 85.0

},

{

“type”: “hasLaterality”,

“sourceId”: “doc1_884540155_M_18”,

“targetId”: “doc1_884540155_M_8”,

“confidence”: 16.5

},

{

“type”: “hasLaterality”,

“sourceId”: “doc1_884540155_M_18”,

“targetId”: “doc1_884540155_M_11”,

“confidence”: 54.599998474121094

},

{

“type”: “hasLaterality”,

“sourceId”: “doc1_884540155_M_18”,

“targetId”: “doc1_884540155_M_12”,

“confidence”: 59.099998474121094

},

{

“type”: “hasLaterality”,

“sourceId”: “doc1_884540155_M_18”,

“targetId”: “doc1_884540155_M_14”,

“confidence”: 30.0

},

{

“type”: “hasLaterality”,

“sourceId”: “doc1_884540155_M_18”,

“targetId”: “doc1_884540155_M_15”,

“confidence”: 49.29999923706055

},

{

“type”: “hasLaterality”,

“sourceId”: “doc1_884540155_M_18”,

“targetId”: “doc1_884540155_M_17”,

“confidence”: 59.95000076293945

},

{

“type”: “hasLaterality”,

“sourceId”: “doc1_884540155_M_18”,

“targetId”: “doc1_884540155_M_30”,

“confidence”: 60.599998474121094

},

{

“type”: “hasLaterality”,

“sourceId”: “doc1_884540155_M_18”,

“targetId”: “doc1_884540155_M_31”,

“confidence”: 52.099998474121094

},

{

“type”: “hasLaterality”,

“sourceId”: “doc1_884540155_M_18”,

“targetId”: “doc1_884540155_M_34”,

“confidence”: 57.099998474121094

},

{

“type”: “hasLaterality”,

“sourceId”: “doc1_884540155_M_18”,

“targetId”: “doc1_884540155_M_37”,

“confidence”: 16.5

},

{

“type”: “hasLaterality”,

“sourceId”: “doc1_884540155_M_18”,

“targetId”: “doc1_884540155_M_41”,

“confidence”: 64.5999984741211

},

{

“type”: “hasLaterality”,

“sourceId”: “doc1_884540155_M_18”,

“targetId”: “doc1_884540155_M_42”,

“confidence”: 24.75

},

{

“type”: “hasLaterality”,

“sourceId”: “doc1_884540155_M_18”,

“targetId”: “doc1_884540155_M_44”,

“confidence”: 26.5

},

{

“type”: “hasQuadrant”,

“sourceId”: “doc1_884540155_M_18”,

“targetId”: “doc1_884540155_M_9”,

“confidence”: 56.79999923706055

},

{

“type”: “hasQuadrant”,

“sourceId”: “doc1_884540155_M_18”,

“targetId”: “doc1_884540155_M_28”,

“confidence”: 54.0

}

]

}

]

},

“neoplasms”: [

{

“id”: “Mass_In_Breast_1683124727155”,

“classUri”: “Mass_In_Breast”,

“attributes”: [

{

“id”: “Upper_inner_Quadrant_1683124727160”,

“classUri”: “Upper_inner_Quadrant”,

“name”: “topography_major”,

“value”: “C502”,

“directEvidence”: [

{

“id”: “doc1_884540155_M_28”,

“classUri”: “Upper_inner_Quadrant”,

“confidence”: 0.0,

“noteId”: “doc1_884540155”,

“begin”: 773,

“end”: 793,

{

“id”: “doc1_884540155_M_45”,

“classUri”: “Breast”,

“confidence”: 0.0,

“noteId”: “doc1_884540155”,

“begin”: 944,

“end”: 950,

“negated”: false,

“uncertain”: false,

“generic”: false,

“conditional”: false,

“historic”: false,

“temporality”: “”

},

{

“id”: “doc1_884540155_M_24”,

“classUri”: “Breast”,

“confidence”: 0.0,

“noteId”: “doc1_884540155”,

“begin”: 1168,

“end”: 1174,

“negated”: false,

“uncertain”: false,

“generic”: false,

“conditional”: false,

“historic”: false,

“temporality”: “”

},

{

“id”: “doc1_884540155_M_9”,

“classUri”: “Upper_inner_Quadrant”,

“confidence”: 0.0,

“noteId”: “doc1_884540155”,

“begin”: 911,

“end”: 931,

“negated”: false,

“uncertain”: false,

“generic”: false,

“conditional”: false,

“historic”: false,

“temporality”: “”

},

{

“id”: “doc1_884540155_M_46”,

“classUri”: “Breast”,

“confidence”: 0.0,

“noteId”: “doc1_884540155”,

“begin”: 545,

“end”: 551,

“negated”: false,

“uncertain”: false,

“generic”: false,

“conditional”: false,

“historic”: false,

“temporality”: “”

},

{

“id”: “doc1_884540155_M_13”,

“classUri”: “Breast”,

“confidence”: 0.0,

“noteId”: “doc1_884540155”,

“begin”: 807,

“end”: 813,

“negated”: false,

“uncertain”: false,

“generic”: false,

“conditional”: false,

“historic”: false,

“temporality”: “”

},

{

“id”: “doc1_884540155_M_26”,

“classUri”: “Breast”,

“confidence”: 0.0,

“noteId”: “doc1_884540155”,

“begin”: 408,

“end”: 415,

“negated”: false,

“uncertain”: false,

“generic”: false,

“conditional”: false,

“historic”: true,

“temporality”: “”

},

{

“id”: “doc1_884540155_M_29”,

“classUri”: “Breast”,

“confidence”: 0.0,

“noteId”: “doc1_884540155”,

“begin”: 2039,

“end”: 2045,

“negated”: false,

“uncertain”: false,

“generic”: false,

“conditional”: false,

“historic”: false,

“temporality”: “”

},

{

“id”: “doc1_884540155_M_47”,

“classUri”: “Breast”,

“confidence”: 0.0,

“noteId”: “doc1_884540155”,

“begin”: 446,

“end”: 452,

“negated”: false,

“uncertain”: false,

“generic”: false,

“conditional”: false,

“historic”: false,

“temporality”: “”

},

{

“id”: “doc1_884540155_M_27”,

“classUri”: “Breast”,

“confidence”: 0.0,

“noteId”: “doc1_884540155”,

“begin”: 1919,

“end”: 1925,

{

“id”: “doc1_884540155_M_10”,

“classUri”: “Breast”,

“confidence”: 0.0,

“noteId”: “doc1_884540155”,

“begin”: 2007,

“end”: 2013,

“negated”: false,

“uncertain”: false,

“generic”: false,

“conditional”: false,

“historic”: false,

“temporality”: “”

},

{

“id”: “doc1_884540155_M_43”,

“classUri”: “Breast”,

“confidence”: 0.0,

“noteId”: “doc1_884540155”,

“begin”: 1068,

“end”: 1074,

“negated”: false,

“uncertain”: false,

“generic”: false,

“conditional”: false,

“historic”: false,

“temporality”: “”

}

],

“indirectEvidence”: [

{

“id”: “doc1_884540155_M_4”,

“classUri”: “Heart”,

“confidence”: 0.0,

“noteId”: “doc1_884540155”,

“begin”: 1765,

“end”: 1772,

“negated”: false,

“uncertain”: false,

“generic”: false,

“conditional”: false,

“historic”: false,

“temporality”: “”

},

{

“id”: “doc1_884540155_M_33”,

“classUri”: “Sclera”,

“confidence”: 0.0,

“noteId”: “doc1_884540155”,

“begin”: 1683,

“end”: 1689,

“negated”: false,

“uncertain”: false,

“generic”: false,

“conditional”: false,

“historic”: false,

“temporality”: “”

},

{

“id”: “doc1_884540155_M_7”,

“classUri”: “Left_Axilla_Proper”,

“confidence”: 0.0,

“noteId”: “doc1_884540155”,

“begin”: 1340,

“end”: 1351,

“negated”: false,

“uncertain”: false,

“generic”: false,

“conditional”: false,

“historic”: false,

“temporality”: “”

},

{

“id”: “doc1_884540155_M_19”,

“classUri”: “Lung”,

“confidence”: 0.0,

“noteId”: “doc1_884540155”,

“begin”: 1726,

“end”: 1731,

“negated”: false,

“uncertain”: false,

“generic”: false,

“conditional”: false,

“historic”: false,

“temporality”: “”

},

{

“id”: “doc1_884540155_M_16”,

“classUri”: “Neck”,

“confidence”: 0.0,

“noteId”: “doc1_884540155”,

“begin”: 1702,

“end”: 1706,

“negated”: false,

“uncertain”: false,

“generic”: false,

“conditional”: false,

“historic”: false,

“temporality”: “”

},

{

“id”: “doc1_884540155_M_40”,

“classUri”: “Ovary”,

“confidence”: 0.0,

“noteId”: “doc1_884540155”,

“begin”: 1432,

“end”: 1439,

“negated”: false,

“uncertain”: false,

“generic”: false,

“conditional”: false,

“historic”: true,

“temporality”: “”

}

],

“notEvidence”: [],

“confidenceFeatures”: []

},

{

“id”: “Upper_Inner_Quadrant_1683124727160”,

“classUri”: “Upper_Inner_Quadrant”,

“name”: “location”,

“value”: “Upper_Inner_Quadrant”,

“directEvidence”: [

{

“classUri”: “Upper_inner_Quadrant”,

“confidence”: 0.0,

“noteId”: “doc1_884540155”,

“begin”: 773,

“end”: 793,

“negated”: false,

“uncertain”: false,

“generic”: false,

“conditional”: false,

“historic”: false,

“temporality”: “”

},

{

“id”: “doc1_884540155_M_45”,

“classUri”: “Breast”,

“confidence”: 0.0,

“noteId”: “doc1_884540155”,

“begin”: 944,

“end”: 950,

“negated”: false,

“uncertain”: false,

“generic”: false,

“conditional”: false,

“historic”: false,

“temporality”: “”

},

{

“id”: “doc1_884540155_M_24”,

“classUri”: “Breast”,

“confidence”: 0.0,

“noteId”: “doc1_884540155”,

“begin”: 1168,

“end”: 1174,

“negated”: false,

“uncertain”: false,

“generic”: false,

“conditional”: false,

“historic”: false,

“temporality”: “”

},

{

“id”: “doc1_884540155_M_9”,

“classUri”: “Upper_inner_Quadrant”,

“confidence”: 0.0,

“noteId”: “doc1_884540155”,

“begin”: 911,

“end”: 931,

“negated”: false,

“uncertain”: false,

“generic”: false,

“conditional”: false,

“historic”: false,

“temporality”: “”

},

{

“id”: “doc1_884540155_M_46”,

“classUri”: “Breast”,

“confidence”: 0.0,

“noteId”: “doc1_884540155”,

“begin”: 545,

“end”: 551,

“negated”: false,

“uncertain”: false,

“generic”: false,

“conditional”: false,

“historic”: false,

“temporality”: “”

},

{

“id”: “doc1_884540155_M_13”,

“classUri”: “Breast”,

“confidence”: 0.0,

“noteId”: “doc1_884540155”,

“begin”: 807,

“end”: 813,

“negated”: false,

“uncertain”: false,

“generic”: false,

“conditional”: false,

“historic”: false,

“temporality”: “”

},

{

“id”: “doc1_884540155_M_26”,

“classUri”: “Breast”,

“confidence”: 0.0,

“noteId”: “doc1_884540155”,

“begin”: 408,

“end”: 415,

“negated”: false,

“uncertain”: false,

“generic”: false,

“conditional”: false,

“historic”: true,

“temporality”: “”

},

{

“id”: “doc1_884540155_M_29”,

“classUri”: “Breast”,

“confidence”: 0.0,

“noteId”: “doc1_884540155”,

“begin”: 2039,

“end”: 2045,

“negated”: false,

“uncertain”: false,

“generic”: false,

“conditional”: false,

“historic”: false,

“temporality”: “”

},

{

“id”: “doc1_884540155_M_47”,

“classUri”: “Breast”,

“confidence”: 0.0,

“noteId”: “doc1_884540155”,

“begin”: 446,

“end”: 452,

“negated”: false,

“uncertain”: false,

“generic”: false,

“conditional”: false,

“historic”: false,

“temporality”: “”

},

{

“classUri”: “Breast”,

“confidence”: 0.0,

“noteId”: “doc1_884540155”,

“begin”: 1919,

“end”: 1925,

“negated”: false,

“uncertain”: false,

“generic”: false,

“conditional”: false,

“historic”: false,

“temporality”: “”

},

{

“id”: “doc1_884540155_M_10”,

“classUri”: “Breast”,

“confidence”: 0.0,

“noteId”: “doc1_884540155”,

“begin”: 2007,

“end”: 2013,

“negated”: false,

“uncertain”: false,

“generic”: false,

“conditional”: false,

“historic”: false,

“temporality”: “”

},

{

“id”: “doc1_884540155_M_43”,

“classUri”: “Breast”,

“confidence”: 0.0,

“noteId”: “doc1_884540155”,

“begin”: 1068,

“end”: 1074,

“negated”: false,

“uncertain”: false,

“generic”: false,

“conditional”: false,

“historic”: false,

“temporality”: “”

}

],

“indirectEvidence”: [

{

“id”: “doc1_884540155_M_4”,

“classUri”: “Heart”,

“confidence”: 0.0,

“noteId”: “doc1_884540155”,

“begin”: 1765,

“end”: 1772,

“negated”: false,

“uncertain”: false,

“generic”: false,

“conditional”: false,

“historic”: false,

“temporality”: “”

},

{

“id”: “doc1_884540155_M_33”,

“classUri”: “Sclera”,

“confidence”: 0.0,

“noteId”: “doc1_884540155”,

“begin”: 1683,

“end”: 1689,

“negated”: false,

“uncertain”: false,

“generic”: false,

“conditional”: false,

“historic”: false,

“temporality”: “”

},

{

“id”: “doc1_884540155_M_7”,

“classUri”: “Left_Axilla_Proper”,

“confidence”: 0.0,

“noteId”: “doc1_884540155”,

“begin”: 1340,

“end”: 1351,

“negated”: false,

“uncertain”: false,

“generic”: false,

“conditional”: false,

“historic”: false,

“temporality”: “”

},

{

“id”: “doc1_884540155_M_19”,

“classUri”: “Lung”,

“confidence”: 0.0,

“noteId”: “doc1_884540155”,

“begin”: 1726,

“end”: 1731,

“negated”: false,

“uncertain”: false,

“generic”: false,

“conditional”: false,

“historic”: false,

“temporality”: “”

},

{

“id”: “doc1_884540155_M_16”,

“classUri”: “Neck”,

“confidence”: 0.0,

“noteId”: “doc1_884540155”,

“begin”: 1702,

“end”: 1706,

“negated”: false,

“uncertain”: false,

“generic”: false,

“conditional”: false,

“historic”: false,

“temporality”: “”

},

{

“id”: “doc1_884540155_M_40”,

“classUri”: “Ovary”,

“confidence”: 0.0,

“noteId”: “doc1_884540155”,

“begin”: 1432,

“end”: 1439,

“negated”: false,

“uncertain”: false,

“generic”: false,

“conditional”: false,

“historic”: true,

“temporality”: “”

}

],

“notEvidence”: [],

“confidenceFeatures”: []

},

{

“id”: “Left_1683124727162”,

“classUri”: “Left”,

“name”: “laterality_code”,

“value”: “2”,

“directEvidence”: [

{

“id”: “doc1_884540155_M_31”,

“classUri”: “Left”,

“confidence”: 0.0,

“noteId”: “doc1_884540155”,

“begin”: 939,

“end”: 943,

“negated”: false,

“uncertain”: false,

“generic”: false,

“conditional”: false,

“historic”: false,

“temporality”: “”

},

{

“id”: “doc1_884540155_M_41”,

“classUri”: “Bilateral”,

“confidence”: 0.0,

“noteId”: “doc1_884540155”,

“begin”: 1494,

“end”: 1503,

“negated”: false,

“uncertain”: false,

“generic”: false,

“conditional”: false,

“historic”: false,

“temporality”: “”

},

{

“id”: “doc1_884540155_M_8”,

“classUri”: “Right”,

“confidence”: 0.0,

“noteId”: “doc1_884540155”,

“begin”: 440,

“end”: 445,

“negated”: false,

“uncertain”: false,

“generic”: false,

“conditional”: false,

“historic”: false,

“temporality”: “”

},

{

“id”: “doc1_884540155_M_17”,

“classUri”: “Left”,

“confidence”: 0.0,

“noteId”: “doc1_884540155”,

“begin”: 1427,

“end”: 1431,

“negated”: false,

“uncertain”: true,

“historic”: true,

“temporality”: “”

},

{

“id”: “doc1_884540155_M_11”,

“classUri”: “Right”,

“confidence”: 0.0,

“noteId”: “doc1_884540155”,

“begin”: 1062,

“end”: 1067,

“negated”: false,

“uncertain”: false,

“generic”: false,

“conditional”: false,

“historic”: false,

“temporality”: “”

},

{

“id”: “doc1_884540155_M_15”,

“classUri”: “Right”,

“confidence”: 0.0,

“noteId”: “doc1_884540155”,

“begin”: 801,

“end”: 806,

“negated”: false,

“uncertain”: false,

“generic”: false,

“conditional”: false,

“historic”: false,

“temporality”: “”

},

{

“id”: “doc1_884540155_M_34”,

“classUri”: “Left”,

“confidence”: 0.0,

“noteId”: “doc1_884540155”,

“begin”: 1163,

“end”: 1167,

“negated”: false,

“uncertain”: false,

“generic”: false,

“conditional”: false,

“historic”: false,

“temporality”: “”

},

{

“id”: “doc1_884540155_M_37”,

“classUri”: “Left”,

“confidence”: 0.0,

“noteId”: “doc1_884540155”,

“begin”: 540,

“end”: 544,

“negated”: false,

“uncertain”: false,

“generic”: false,

“conditional”: false,

“historic”: false,

“temporality”: “”

},

{

“id”: “doc1_884540155_M_14”,

“classUri”: “Bilateral”,

“confidence”: 0.0,

“noteId”: “doc1_884540155”,

“begin”: 1909,

“end”: 1918,

“negated”: false,

“uncertain”: false,

“generic”: false,

“conditional”: false,

“historic”: false,

“temporality”: “”

},

{

“id”: “doc1_884540155_M_30”,

“classUri”: “Left”,

“confidence”: 0.0,

“noteId”: “doc1_884540155”,

“begin”: 1340,

“end”: 1344,

“negated”: false,

“uncertain”: false,

“generic”: false,

“conditional”: false,

“historic”: false,

“temporality”: “”

},

{

“id”: “doc1_884540155_M_44”,

“classUri”: “Right”,

“confidence”: 0.0,

“noteId”: “doc1_884540155”,

“begin”: 2001,

“end”: 2006,

“negated”: false,

“uncertain”: false,

“generic”: false,

“conditional”: false,

“historic”: false,

“temporality”: “”

},

{

“id”: “doc1_884540155_M_12”,

“classUri”: “Right”,

“confidence”: 0.0,

“noteId”: “doc1_884540155”,

“begin”: 1240,

“end”: 1245,

“negated”: false,

“uncertain”: false,

“generic”: false,

“conditional”: false,

“historic”: false,

“temporality”: “”

},

{

“id”: “doc1_884540155_M_42”,

“classUri”: “Left”,

“confidence”: 0.0,

“noteId”: “doc1_884540155”,

“begin”: 2034,

“end”: 2038,

“negated”: false,

“uncertain”: false,

“historic”: false,

“temporality”: “”

}

],

“indirectEvidence”: [],

“notEvidence”: [],

“confidenceFeatures”: []

},

{

“id”: “Left_1683124727162”,

“classUri”: “Left”,

“name”: “laterality”,

“value”: “Left”,

“directEvidence”: [

{

“id”: “doc1_884540155_M_31”,

“classUri”: “Left”,

“confidence”: 0.0,

“noteId”: “doc1_884540155”,

“begin”: 939,

“end”: 943,

“negated”: false,

“uncertain”: false,

“generic”: false,

“conditional”: false,

“historic”: false,

“temporality”: “”

},

{

“id”: “doc1_884540155_M_41”,

“classUri”: “Bilateral”,

“confidence”: 0.0,

“noteId”: “doc1_884540155”,

“begin”: 1494,

“end”: 1503,

“negated”: false,

“uncertain”: false,

“generic”: false,

“conditional”: false,

“historic”: false,

“temporality”: “”

},

{

“id”: “doc1_884540155_M_8”,

“classUri”: “Right”,

“confidence”: 0.0,

“noteId”: “doc1_884540155”,

“begin”: 440,

“end”: 445,

“negated”: false,

“uncertain”: false,

“generic”: false,

“conditional”: false,

“historic”: false,

“temporality”: “”

},

{

“id”: “doc1_884540155_M_17”,

“classUri”: “Left”,

“confidence”: 0.0,

“noteId”: “doc1_884540155”,

“begin”: 1427,

“negated”: false,

“uncertain”: true,

“generic”: false,

“conditional”: false,

“historic”: true,

“temporality”: “”

},

{

“id”: “doc1_884540155_M_11”,

“classUri”: “Right”,

“confidence”: 0.0,

“noteId”: “doc1_884540155”,

“begin”: 1062,

“end”: 1067,

“negated”: false,

“uncertain”: false,

“generic”: false,

“conditional”: false,

“historic”: false,

“temporality”: “”

},

{

“id”: “doc1_884540155_M_15”,

“classUri”: “Right”,

“confidence”: 0.0,

“noteId”: “doc1_884540155”,

“begin”: 801,

“end”: 806,

“negated”: false,

“uncertain”: false,

“generic”: false,

“conditional”: false,

“historic”: false,

“temporality”: “”

},

{

“id”: “doc1_884540155_M_34”,

“classUri”: “Left”,

“confidence”: 0.0,

“noteId”: “doc1_884540155”,

“begin”: 1163,

“end”: 1167,

“negated”: false,

“uncertain”: false,

“generic”: false,

“conditional”: false,

“historic”: false,

“temporality”: “”

},

{

“id”: “doc1_884540155_M_37”,

“classUri”: “Left”,

“confidence”: 0.0,

“noteId”: “doc1_884540155”,

“begin”: 540,

“end”: 544,

“negated”: false,

“uncertain”: false,

“generic”: false,

“conditional”: false,

“historic”: false,

“temporality”: “”

},

{

“id”: “doc1_884540155_M_14”,

“classUri”: “Bilateral”,

“confidence”: 0.0,

“noteId”: “doc1_884540155”,

“begin”: 1909,

“end”: 1918,

“negated”: false,

“uncertain”: false,

“generic”: false,

“conditional”: false,

“historic”: false,

“temporality”: “”

},

{

“id”: “doc1_884540155_M_30”,

“classUri”: “Left”,

“confidence”: 0.0,

“noteId”: “doc1_884540155”,

“begin”: 1340,

“end”: 1344,

“negated”: false,

“uncertain”: false,

“generic”: false,

“conditional”: false,

“historic”: false,

“temporality”: “”

},

{

“id”: “doc1_884540155_M_44”,

“classUri”: “Right”,

“confidence”: 0.0,

“noteId”: “doc1_884540155”,

“begin”: 2001,

“end”: 2006,

“negated”: false,

“uncertain”: false,

“generic”: false,

“conditional”: false,

“historic”: false,

“temporality”: “”

},

{

“id”: “doc1_884540155_M_12”,

“classUri”: “Right”,

“confidence”: 0.0,

“noteId”: “doc1_884540155”,

“begin”: 1240,

“end”: 1245,

“negated”: false,

“uncertain”: false,

“generic”: false,

“conditional”: false,

“historic”: false,

“temporality”: “”

},

{

“id”: “doc1_884540155_M_42”,

“classUri”: “Left”,

“confidence”: 0.0,

“noteId”: “doc1_884540155”,

“begin”: 2034,

“negated”: false,

“uncertain”: false,

“generic”: false,

“conditional”: false,

“historic”: false,

“temporality”: “”

}

],

“indirectEvidence”: [],

“notEvidence”: [],

“confidenceFeatures”: []

},

{

“id”: “Upper_inner_Quadrant_1683124727168”,

“classUri”: “Upper_inner_Quadrant”,

“name”: “topography_minor”,

“value”: “9”,

“directEvidence”: [

{

“id”: “doc1_884540155_M_45”,

“classUri”: “Breast”,

“confidence”: 0.0,

“noteId”: “doc1_884540155”,

“begin”: 944,

“end”: 950,

“negated”: false,

“uncertain”: false,

“generic”: false,

“conditional”: false,

“historic”: false,

“temporality”: “”

},

{

“id”: “doc1_884540155_M_24”,

“classUri”: “Breast”,

“confidence”: 0.0,

“noteId”: “doc1_884540155”,

“begin”: 1168,

“end”: 1174,

“negated”: false,

“uncertain”: false,

“generic”: false,

“conditional”: false,

“historic”: false,

“temporality”: “”

},

{

“id”: “doc1_884540155_M_46”,

“classUri”: “Breast”,

“confidence”: 0.0,

“noteId”: “doc1_884540155”,

“begin”: 545,

“end”: 551,

“negated”: false,

“uncertain”: false,

“generic”: false,

“conditional”: false,

“historic”: false,

“temporality”: “”

},

{

“confidence”: 0.0,

“noteId”: “doc1_884540155”,

“begin”: 446,

“end”: 452,

“negated”: false,

“uncertain”: false,

“generic”: false,

“conditional”: false,

“historic”: false,

“temporality”: “”

},

{

“id”: “doc1_884540155_M_27”,

“classUri”: “Breast”,

“confidence”: 0.0,

“noteId”: “doc1_884540155”,

“begin”: 1919,

“end”: 1925,

“negated”: false,

“uncertain”: false,

“generic”: false,

“conditional”: false,

“historic”: false,

“temporality”: “”

},

{

“id”: “doc1_884540155_M_19”,

“classUri”: “Lung”,

“confidence”: 0.0,

“noteId”: “doc1_884540155”,

“begin”: 1726,

“end”: 1731,

“negated”: false,

“uncertain”: false,

“generic”: false,

“conditional”: false,

“historic”: false,

“temporality”: “”

},

{

“id”: “doc1_884540155_M_43”,

“classUri”: “Breast”,

“confidence”: 0.0,

“noteId”: “doc1_884540155”,

“begin”: 1068,

“end”: 1074,

“negated”: false,

“uncertain”: false,

“generic”: false,

“conditional”: false,

“historic”: false,

“temporality”: “”

},

{

“id”: “doc1_884540155_M_28”,

“classUri”: “Upper_inner_Quadrant”,

“confidence”: 0.0,

“noteId”: “doc1_884540155”,

“begin”: 773,

“end”: 793,

“negated”: false,

“uncertain”: false,

“generic”: false,

“conditional”: false,

“historic”: false,

“temporality”: “”

},

{

“id”: “doc1_884540155_M_9”,

“classUri”: “Upper_inner_Quadrant”,

“confidence”: 0.0,

“noteId”: “doc1_884540155”,

“begin”: 911,

“end”: 931,

“negated”: false,

“uncertain”: false,

“generic”: false,

“conditional”: false,

“historic”: false,

“temporality”: “”

},

{

“id”: “doc1_884540155_M_13”,

“classUri”: “Breast”,

“confidence”: 0.0,

“noteId”: “doc1_884540155”,

“begin”: 807,

“end”: 813,

“negated”: false,

“uncertain”: false,

“generic”: false,

“conditional”: false,

“historic”: false,

“temporality”: “”

},

{

“id”: “doc1_884540155_M_26”,

“classUri”: “Breast”,

“confidence”: 0.0,

“noteId”: “doc1_884540155”,

“begin”: 408,

“end”: 415,

“negated”: false,

“uncertain”: false,

“generic”: false,

“conditional”: false,

“historic”: true,

“temporality”: “”

},

{

“id”: “doc1_884540155_M_29”,

“classUri”: “Breast”,

“confidence”: 0.0,

“noteId”: “doc1_884540155”,

“begin”: 2039,

“end”: 2045,

“negated”: false,

“uncertain”: false,

“generic”: false,

“conditional”: false,

“historic”: false,

“temporality”: “”

},

{

“confidence”: 0.0,

“noteId”: “doc1_884540155”,

“begin”: 2007,

“end”: 2013,

“negated”: false,

“uncertain”: false,

“generic”: false,

“conditional”: false,

“historic”: false,

“temporality”: “”

}

],

“indirectEvidence”: [],

“notEvidence”: [],

“confidenceFeatures”: []

},

{

“id”: “Breast_Quadrant_1683124727168”,

“classUri”: “Breast_Quadrant”,

“name”: “quadrant”,

“value”: “Upper_inner_Quadrant”,

“directEvidence”: [

{

“id”: “doc1_884540155_M_45”,

“classUri”: “Breast”,

“confidence”: 0.0,

“noteId”: “doc1_884540155”,

“begin”: 944,

“end”: 950,

“negated”: false,

“uncertain”: false,

“generic”: false,

“conditional”: false,

“historic”: false,

“temporality”: “”

},

{

“id”: “doc1_884540155_M_24”,

“classUri”: “Breast”,

“confidence”: 0.0,

“noteId”: “doc1_884540155”,

“begin”: 1168,

“end”: 1174,

“negated”: false,

“uncertain”: false,

“generic”: false,

“conditional”: false,

“historic”: false,

“temporality”: “”

},

{

“id”: “doc1_884540155_M_46”,

“classUri”: “Breast”,

“confidence”: 0.0,

“noteId”: “doc1_884540155”,

“begin”: 545,

“end”: 551,

},

{

“id”: “doc1_884540155_M_47”,

“classUri”: “Breast”,

“confidence”: 0.0,

“noteId”: “doc1_884540155”,

“begin”: 446,

“end”: 452,

“negated”: false,

“uncertain”: false,

“generic”: false,

“conditional”: false,

“historic”: false,

“temporality”: “”

},

{

“id”: “doc1_884540155_M_27”,

“classUri”: “Breast”,

“confidence”: 0.0,

“noteId”: “doc1_884540155”,

“begin”: 1919,

“end”: 1925,

“negated”: false,

“uncertain”: false,

“generic”: false,

“conditional”: false,

“historic”: false,

“temporality”: “”

},

{

“id”: “doc1_884540155_M_19”,

“classUri”: “Lung”,

“confidence”: 0.0,

“noteId”: “doc1_884540155”,

“begin”: 1726,

“end”: 1731,

“negated”: false,

“uncertain”: false,

“generic”: false,

“conditional”: false,

“historic”: false,

“temporality”: “”

},

{

“id”: “doc1_884540155_M_43”,

“classUri”: “Breast”,

“confidence”: 0.0,

“noteId”: “doc1_884540155”,

“begin”: 1068,

“end”: 1074,

“negated”: false,

“uncertain”: false,

“generic”: false,

“conditional”: false,

“historic”: false,

“temporality”: “”

},

{

“id”: “doc1_884540155_M_28”,

“classUri”: “Upper_inner_Quadrant”,

“confidence”: 0.0,

“noteId”: “doc1_884540155”,

“begin”: 773,

“end”: 793,

“negated”: false,

“uncertain”: false,

“generic”: false,

“conditional”: false,

“historic”: false,

“temporality”: “”

},

{

“id”: “doc1_884540155_M_9”,

“classUri”: “Upper_inner_Quadrant”,

“confidence”: 0.0,

“noteId”: “doc1_884540155”,

“begin”: 911,

“end”: 931,

“negated”: false,

“uncertain”: false,

“generic”: false,

“conditional”: false,

“historic”: false,

“temporality”: “”

},

{

“id”: “doc1_884540155_M_13”,

“classUri”: “Breast”,

“confidence”: 0.0,

“noteId”: “doc1_884540155”,

“begin”: 807,

“end”: 813,

“negated”: false,

“uncertain”: false,

“generic”: false,

“conditional”: false,

“historic”: false,

“temporality”: “”

},

{

“id”: “doc1_884540155_M_26”,

“classUri”: “Breast”,

“confidence”: 0.0,

“noteId”: “doc1_884540155”,

“begin”: 408,

“end”: 415,

“negated”: false,

“uncertain”: false,

“generic”: false,

“conditional”: false,

“historic”: true,

“temporality”: “”

},

{

“id”: “doc1_884540155_M_29”,

“classUri”: “Breast”,

“confidence”: 0.0,

“noteId”: “doc1_884540155”,

“begin”: 2039,

“end”: 2045,

},

{

“id”: “doc1_884540155_M_10”,

“classUri”: “Breast”,

“confidence”: 0.0,

“noteId”: “doc1_884540155”,

“begin”: 2007,

“end”: 2013,

“negated”: false,

“uncertain”: false,

“generic”: false,

“conditional”: false,

“historic”: false,

“temporality”: “”

}

],

“indirectEvidence”: [],

“notEvidence”: [],

“confidenceFeatures”: []

},

{

“id”: “Mass_In_Breast_1683124727175”,

“classUri”: “Mass_In_Breast”,

“name”: “histology”,

“value”: “8727”,

“directEvidence”: [

{

“id”: “doc1_884540155_M_22”,

“classUri”: “Mass”,

“confidence”: 0.0,

“noteId”: “doc1_884540155”,

“begin”: 528,

“end”: 532,

“negated”: false,

“uncertain”: false,

“generic”: false,

“conditional”: false,

“historic”: false,

“temporality”: “”

},

{

“id”: “doc1_884540155_M_3”,

“classUri”: “Mass_In_Breast”,

“confidence”: 0.0,

“noteId”: “doc1_884540155”,

“begin”: 402,

“end”: 415,

“negated”: false,

“uncertain”: true,

“generic”: false,

“conditional”: false,

“historic”: true,

“temporality”: “”

},

{

“id”: “doc1_884540155_M_23”,

“classUri”: “Lobular_Mass”,

“confidence”: 0.0,

“noteId”: “doc1_884540155”,

“begin”: 751,

“end”: 765,

“negated”: false,

“uncertain”: false,

“generic”: false,

“conditional”: false,

“historic”: false,

“temporality”: “”

},

{

“id”: “doc1_884540155_M_20”,

“classUri”: “Dysplastic_Nevus”,

“confidence”: 0.0,

“noteId”: “doc1_884540155”,

“begin”: 1650,

“end”: 1653,

“negated”: false,

“uncertain”: false,

“generic”: false,

“conditional”: false,

“historic”: false,

“temporality”: “”

},

{

“id”: “doc1_884540155_M_35”,

“classUri”: “Mass_In_Breast”,

“confidence”: 0.0,

“noteId”: “doc1_884540155”,

“begin”: 2039,

“end”: 2052,

“negated”: false,

“uncertain”: false,

“generic”: false,

“conditional”: false,

“historic”: false,

“temporality”: “”

},

{

“id”: “doc1_884540155_M_36”,

“classUri”: “Mass”,

“confidence”: 0.0,

“noteId”: “doc1_884540155”,

“begin”: 899,

“end”: 903,

“negated”: false,

“uncertain”: false,

“generic”: false,

“conditional”: false,

“historic”: false,

“temporality”: “”

},

{

“id”: “doc1_884540155_M_5”,

“classUri”: “Mass_In_Breast”,

“confidence”: 0.0,

“noteId”: “doc1_884540155”,

“begin”: 2007,

“end”: 2018,

“negated”: false,

“uncertain”: false,

“generic”: false,

“conditional”: false,

“historic”: false,

“temporality”: “”

},

{

“id”: “doc1_884540155_M_38”,

“classUri”: “Mass”,

“confidence”: 0.0,

“noteId”: “doc1_884540155”,

“begin”: 1107,

“end”: 1111,

“negated”: false,

“uncertain”: false,

“generic”: false,

“conditional”: false,

“historic”: false,

“temporality”: “”

},

{

“id”: “doc1_884540155_M_21”,

“classUri”: “Mass”,

“confidence”: 0.0,

“noteId”: “doc1_884540155”,

“begin”: 428,

“end”: 432,

“negated”: false,

“uncertain”: false,

“generic”: false,

“conditional”: false,

“historic”: false,

“temporality”: “”

},

{

“id”: “doc1_884540155_M_18”,

“classUri”: “Epithelial_Ovarian_Cancer”,

“confidence”: 0.0,

“noteId”: “doc1_884540155”,

“begin”: 1432,

“end”: 1449,

“negated”: false,

“uncertain”: false,

“generic”: false,

“conditional”: false,

“historic”: true,

“temporality”: “”

},

{

“id”: “doc1_884540155_M_2”,

“classUri”: “Mass”,

“confidence”: 0.0,

“noteId”: “doc1_884540155”,

“begin”: 1215,

“end”: 1219,

“negated”: false,

“uncertain”: false,

“generic”: false,

“conditional”: false,

“historic”: false,

“temporality”: “”

}

],

“indirectEvidence”: [],

“notEvidence”: [],

“confidenceFeatures”: []

},

{

“id”: “Mass_In_Breast_1683124727175”,

“classUri”: “Mass_In_Breast”,

“name”: “cancer_type”,

“value”: “Mass_In_Breast”,

“directEvidence”: [

{

“id”: “doc1_884540155_M_22”,

“classUri”: “Mass”,

“confidence”: 0.0,

“noteId”: “doc1_884540155”,

“begin”: 528,

“end”: 532,

“negated”: false,

“uncertain”: false,

“generic”: false,

“conditional”: false,

“historic”: false,

“temporality”: “”

},

{

“id”: “doc1_884540155_M_3”,

“classUri”: “Mass_In_Breast”,

“confidence”: 0.0,

“noteId”: “doc1_884540155”,

“begin”: 402,

“end”: 415,

“negated”: false,

“uncertain”: true,

“generic”: false,

“conditional”: false,

“historic”: true,

“temporality”: “”

},

{

“id”: “doc1_884540155_M_23”,

“classUri”: “Lobular_Mass”,

“confidence”: 0.0,

“noteId”: “doc1_884540155”,

“begin”: 751,

“end”: 765,

“negated”: false,

“uncertain”: false,

“generic”: false,

“conditional”: false,

“historic”: false,

“temporality”: “”

},

{

“id”: “doc1_884540155_M_20”,

“classUri”: “Dysplastic_Nevus”,

“confidence”: 0.0,

“noteId”: “doc1_884540155”,

“begin”: 1650,

“end”: 1653,

“negated”: false,

“uncertain”: false,

“generic”: false,

“conditional”: false,

“historic”: false,

“temporality”: “”

},

{

“id”: “doc1_884540155_M_35”,

“classUri”: “Mass_In_Breast”,

“confidence”: 0.0,

“noteId”: “doc1_884540155”,

“begin”: 2039,

“end”: 2052,

“negated”: false,

“uncertain”: false,

“generic”: false,

“conditional”: false,

“historic”: false,

“temporality”: “”

},

{

“id”: “doc1_884540155_M_36”,

“classUri”: “Mass”,

“confidence”: 0.0,

“noteId”: “doc1_884540155”,

“begin”: 899,

“end”: 903,

“negated”: false,

“uncertain”: false,

“generic”: false,

“conditional”: false,

“historic”: false,

“temporality”: “”

},

{

“id”: “doc1_884540155_M_5”,

“classUri”: “Mass_In_Breast”,

“confidence”: 0.0,

“noteId”: “doc1_884540155”,

“begin”: 2007,

“end”: 2018,

“negated”: false,

“uncertain”: false,

“generic”: false,

“conditional”: false,

“historic”: false,

“temporality”: “”

},

{

“id”: “doc1_884540155_M_38”,

“classUri”: “Mass”,

“confidence”: 0.0,

“noteId”: “doc1_884540155”,

“begin”: 1107,

“end”: 1111,

“negated”: false,

“uncertain”: false,

“generic”: false,

“conditional”: false,

“historic”: false,

“temporality”: “”

},

{

“id”: “doc1_884540155_M_21”,

“classUri”: “Mass”,

“confidence”: 0.0,

“noteId”: “doc1_884540155”,

“begin”: 428,

“end”: 432,

“negated”: false,

“uncertain”: false,

“generic”: false,

“conditional”: false,

“historic”: false,

“temporality”: “”

},

{

“id”: “doc1_884540155_M_18”,

“classUri”: “Epithelial_Ovarian_Cancer”,

“confidence”: 0.0,

“noteId”: “doc1_884540155”,

“begin”: 1432,

“end”: 1449,

“negated”: false,

“uncertain”: false,

“generic”: false,

“conditional”: false,

“historic”: true,

“temporality”: “”

},

{

“id”: “doc1_884540155_M_2”,

“classUri”: “Mass”,

“confidence”: 0.0,

“noteId”: “doc1_884540155”,

“begin”: 1215,

“end”: 1219,

“negated”: false,

“uncertain”: false,

“generic”: false,

“conditional”: false,

“historic”: false,

“temporality”: “”

}

],

“indirectEvidence”: [],

“notEvidence”: [],

“confidenceFeatures”: []

},

{

“id”: “Mass_In_Breast_1683124727175”,

“classUri”: “Mass_In_Breast”,

“name”: “histologic_type”,

“value”: “Mass_In_Breast”,

“directEvidence”: [

{

“id”: “doc1_884540155_M_22”,

“classUri”: “Mass”,

“confidence”: 0.0,

“noteId”: “doc1_884540155”,

“begin”: 528,

“end”: 532,

“negated”: false,

“uncertain”: false,

“generic”: false,

“conditional”: false,

“historic”: false,

“temporality”: “”

},

{

“id”: “doc1_884540155_M_3”,

“classUri”: “Mass_In_Breast”,

“confidence”: 0.0,

“noteId”: “doc1_884540155”,

“begin”: 402,

“end”: 415,

“uncertain”: true,

“generic”: false,

“conditional”: false,

“historic”: true,

“temporality”: “”

},

{

“id”: “doc1_884540155_M_23”,

“classUri”: “Lobular_Mass”,

“confidence”: 0.0,

“noteId”: “doc1_884540155”,

“begin”: 751,

“end”: 765,

“negated”: false,

“uncertain”: false,

“generic”: false,

“conditional”: false,

“historic”: false,

“temporality”: “”

},

{

“id”: “doc1_884540155_M_20”,

“classUri”: “Dysplastic_Nevus”,

“confidence”: 0.0,

“noteId”: “doc1_884540155”,

“begin”: 1650,

“end”: 1653,

“negated”: false,

“uncertain”: false,

“generic”: false,

“conditional”: false,

“historic”: false,

“temporality”: “”

},

{

“id”: “doc1_884540155_M_35”,

“classUri”: “Mass_In_Breast”,

“confidence”: 0.0,

“noteId”: “doc1_884540155”,

“begin”: 2039,

“end”: 2052,

“negated”: false,

“uncertain”: false,

“generic”: false,

“conditional”: false,

“historic”: false,

“temporality”: “”

},

{

“id”: “doc1_884540155_M_36”,

“classUri”: “Mass”,

“confidence”: 0.0,

“noteId”: “doc1_884540155”,

“begin”: 899,

“end”: 903,

“negated”: false,

“uncertain”: false,

“generic”: false,

“conditional”: false,

“historic”: false,

“temporality”: “”

},

{

“id”: “doc1_884540155_M_5”,

“classUri”: “Mass_In_Breast”,

“confidence”: 0.0,

“noteId”: “doc1_884540155”,

“begin”: 2007,

“end”: 2018,

“negated”: false,

“uncertain”: false,

“generic”: false,

“conditional”: false,

“historic”: false,

“temporality”: “”

},

{

“id”: “doc1_884540155_M_38”,

“classUri”: “Mass”,

“confidence”: 0.0,

“noteId”: “doc1_884540155”,

“begin”: 1107,

“end”: 1111,

“negated”: false,

“uncertain”: false,

“generic”: false,

“conditional”: false,

“historic”: false,

“temporality”: “”

},

{

“id”: “doc1_884540155_M_21”,

“classUri”: “Mass”,

“confidence”: 0.0,

“noteId”: “doc1_884540155”,

“begin”: 428,

“end”: 432,

“negated”: false,

“uncertain”: false,

“generic”: false,

“conditional”: false,

“historic”: false,

“temporality”: “”

},

{

“id”: “doc1_884540155_M_18”,

“classUri”: “Epithelial_Ovarian_Cancer”,

“confidence”: 0.0,

“noteId”: “doc1_884540155”,

“begin”: 1432,

“end”: 1449,

“negated”: false,

“uncertain”: false,

“generic”: false,

“conditional”: false,

“historic”: true,

“temporality”: “”

},

{

“id”: “doc1_884540155_M_2”,

“classUri”: “Mass”,

“confidence”: 0.0,

“noteId”: “doc1_884540155”,

“begin”: 1215,

“end”: 1219,

“uncertain”: false,

“generic”: false,

“conditional”: false,

“historic”: false,

“temporality”: “”

}

],

“indirectEvidence”: [],

“notEvidence”: [],

“confidenceFeatures”: []

},

{

“id”: “Mass_In_Breast_1683124727175”,

“classUri”: “Mass_In_Breast”,

“name”: “diagnosis”,

“value”: “Mass_In_Breast”,

“directEvidence”: [

{

“id”: “doc1_884540155_M_22”,

“classUri”: “Mass”,

“confidence”: 0.0,

“noteId”: “doc1_884540155”,

“begin”: 528,

“end”: 532,

“negated”: false,

“uncertain”: false,

“generic”: false,

“conditional”: false,

“historic”: false,

“temporality”: “”

},

{

“id”: “doc1_884540155_M_3”,

“classUri”: “Mass_In_Breast”,

“confidence”: 0.0,

“noteId”: “doc1_884540155”,

“begin”: 402,

“end”: 415,

“negated”: false,

“uncertain”: true,

“generic”: false,

“conditional”: false,

“historic”: true,

“temporality”: “”

},

{

“id”: “doc1_884540155_M_23”,

“classUri”: “Lobular_Mass”,

“confidence”: 0.0,

“noteId”: “doc1_884540155”,

“begin”: 751,

“end”: 765,

“negated”: false,

“uncertain”: false,

“generic”: false,

“conditional”: false,

“historic”: false,

“temporality”: “”

},

{

“id”: “doc1_884540155_M_20”,

“classUri”: “Dysplastic_Nevus”,

“confidence”: 0.0,

“noteId”: “doc1_884540155”,

“begin”: 1650,

“end”: 1653,

“negated”: false,

“uncertain”: false,

“generic”: false,

“conditional”: false,

“historic”: false,

“temporality”: “”

},

{

“id”: “doc1_884540155_M_35”,

“classUri”: “Mass_In_Breast”,

“confidence”: 0.0,

“noteId”: “doc1_884540155”,

“begin”: 2039,

“end”: 2052,

“negated”: false,

“uncertain”: false,

“generic”: false,

“conditional”: false,

“historic”: false,

“temporality”: “”

},

{

“id”: “doc1_884540155_M_36”,

“classUri”: “Mass”,

“confidence”: 0.0,

“noteId”: “doc1_884540155”,

“begin”: 899,

“end”: 903,

“negated”: false,

“uncertain”: false,

“generic”: false,

“conditional”: false,

“historic”: false,

“temporality”: “”

},

{

“id”: “doc1_884540155_M_5”,

“classUri”: “Mass_In_Breast”,

“confidence”: 0.0,

“noteId”: “doc1_884540155”,

“begin”: 2007,

“end”: 2018,

“negated”: false,

“uncertain”: false,

“generic”: false,

“conditional”: false,

“historic”: false,

“temporality”: “”

},

{

“id”: “doc1_884540155_M_38”,

“classUri”: “Mass”,

“confidence”: 0.0,

“noteId”: “doc1_884540155”,

“begin”: 1107,

“end”: 1111,

“negated”: false,

“uncertain”: false,

“generic”: false,

“conditional”: false,

“historic”: false,

“temporality”: “”

},

{

“id”: “doc1_884540155_M_21”,

“classUri”: “Mass”,

“confidence”: 0.0,

“noteId”: “doc1_884540155”,

“begin”: 428,

“end”: 432,

“negated”: false,

“uncertain”: false,

“generic”: false,

“conditional”: false,

“historic”: false,

“temporality”: “”

},

{

“id”: “doc1_884540155_M_18”,

“classUri”: “Epithelial_Ovarian_Cancer”,

“confidence”: 0.0,

“noteId”: “doc1_884540155”,

“begin”: 1432,

“end”: 1449,

“negated”: false,

“uncertain”: false,

“generic”: false,

“conditional”: false,

“historic”: true,

“temporality”: “”

},

{

“id”: “doc1_884540155_M_2”,

“classUri”: “Mass”,

“confidence”: 0.0,

“noteId”: “doc1_884540155”,

“begin”: 1215,

“end”: 1219,

“negated”: false,

“uncertain”: false,

“generic”: false,

“conditional”: false,

“historic”: false,

“temporality”: “”

}

],

“indirectEvidence”: [],

“notEvidence”: [],

“confidenceFeatures”: []

},

{

“id”: “Mass_In_Breast_1683124727183”,

“classUri”: “Mass_In_Breast”,

“name”: “behavior”,

“value”: “3”,

“directEvidence”: [

{

“id”: “doc1_884540155_M_22”,

“classUri”: “Mass”,

“confidence”: 0.0,

“noteId”: “doc1_884540155”,

“begin”: 528,

“end”: 532,

“negated”: false,

“uncertain”: false,

“generic”: false,

“conditional”: false,

“historic”: false,

“temporality”: “”

},

{

“id”: “doc1_884540155_M_3”,

“classUri”: “Mass_In_Breast”,

“confidence”: 0.0,

“noteId”: “doc1_884540155”,

“begin”: 402,

“end”: 415,

“negated”: false,

“uncertain”: true,

“generic”: false,

“conditional”: false,

“historic”: true,

“temporality”: “”

},

{

“id”: “doc1_884540155_M_23”,

“classUri”: “Lobular_Mass”,

“confidence”: 0.0,

“noteId”: “doc1_884540155”,

“begin”: 751,

“end”: 765,

“negated”: false,

“uncertain”: false,

“generic”: false,

“conditional”: false,

“historic”: false,

“temporality”: “”

},

{

“id”: “doc1_884540155_M_20”,

“classUri”: “Dysplastic_Nevus”,

“confidence”: 0.0,

“noteId”: “doc1_884540155”,

“begin”: 1650,

“end”: 1653,

“negated”: false,

“uncertain”: false,

“generic”: false,

“conditional”: false,

“historic”: false,

“temporality”: “”

},

{

“id”: “doc1_884540155_M_35”,

“classUri”: “Mass_In_Breast”,

“confidence”: 0.0,

“noteId”: “doc1_884540155”,

“begin”: 2039,

“end”: 2052,

“negated”: false,

“uncertain”: false,

“generic”: false,

“conditional”: false,

“historic”: false,

“temporality”: “”

},

{

“id”: “doc1_884540155_M_36”,

“classUri”: “Mass”,

“confidence”: 0.0,

“noteId”: “doc1_884540155”,

“begin”: 899,

“end”: 903,

“negated”: false,

“uncertain”: false,

“generic”: false,

“conditional”: false,

“historic”: false,

“temporality”: “”

},

{

“id”: “doc1_884540155_M_5”,

“classUri”: “Mass_In_Breast”,

“confidence”: 0.0,

“noteId”: “doc1_884540155”,

“begin”: 2007,

“end”: 2018,

“negated”: false,

“uncertain”: false,

“generic”: false,

“conditional”: false,

“historic”: false,

“temporality”: “”

},

{

“id”: “doc1_884540155_M_38”,

“classUri”: “Mass”,

“confidence”: 0.0,

“noteId”: “doc1_884540155”,

“begin”: 1107,

“end”: 1111,

“negated”: false,

“uncertain”: false,

“generic”: false,

“conditional”: false,

“historic”: false,

“temporality”: “”

},

{

“id”: “doc1_884540155_M_21”,

“classUri”: “Mass”,

“confidence”: 0.0,

“noteId”: “doc1_884540155”,

“begin”: 428,

“end”: 432,

“negated”: false,

“uncertain”: false,

“generic”: false,

“conditional”: false,

“historic”: false,

“temporality”: “”

},

{

“id”: “doc1_884540155_M_18”,

“classUri”: “Epithelial_Ovarian_Cancer”,

“confidence”: 0.0,

“noteId”: “doc1_884540155”,

“begin”: 1432,

“end”: 1449,

“negated”: false,

“uncertain”: false,

“generic”: false,

“conditional”: false,

“historic”: true,

“temporality”: “”

},

{

“id”: “doc1_884540155_M_2”,

“classUri”: “Mass”,

“confidence”: 0.0,

“noteId”: “doc1_884540155”,

“begin”: 1215,

“end”: 1219,

“negated”: false,

“uncertain”: false,

“generic”: false,

“conditional”: false,

“historic”: false,

“temporality”: “”

}

],

“indirectEvidence”: [],

“notEvidence”: [],

“confidenceFeatures”: []

},

{

“id”: “Mass_In_Breast_1683124727183”,

“classUri”: “Mass_In_Breast”,

“name”: “extent”,

“value”: “Invasive_Lesion”,

“directEvidence”: [

{

“id”: “doc1_884540155_M_22”,

“classUri”: “Mass”,

“confidence”: 0.0,

“noteId”: “doc1_884540155”,

“begin”: 528,

“end”: 532,

“negated”: false,

“uncertain”: false,

“generic”: false,

“conditional”: false,

“historic”: false,

“temporality”: “”

},

{

“id”: “doc1_884540155_M_3”,

“classUri”: “Mass_In_Breast”,

“confidence”: 0.0,

“noteId”: “doc1_884540155”,

“begin”: 402,

“end”: 415,

“negated”: false,

“uncertain”: true,

“generic”: false,

“conditional”: false,

“historic”: true,

“temporality”: “”

},

{

“id”: “doc1_884540155_M_23”,

“classUri”: “Lobular_Mass”,

“confidence”: 0.0,

“noteId”: “doc1_884540155”,

“begin”: 751,

“end”: 765,

“negated”: false,

“uncertain”: false,

“generic”: false,

“conditional”: false,

“historic”: false,

“temporality”: “”

},

{

“id”: “doc1_884540155_M_20”,

“classUri”: “Dysplastic_Nevus”,

“confidence”: 0.0,

“noteId”: “doc1_884540155”,

“begin”: 1650,

“end”: 1653,

“negated”: false,

“uncertain”: false,

“generic”: false,

“conditional”: false,

“historic”: false,

“temporality”: “”

},

{

“id”: “doc1_884540155_M_35”,

“classUri”: “Mass_In_Breast”,

“confidence”: 0.0,

“noteId”: “doc1_884540155”,

“begin”: 2039,

“end”: 2052,

“negated”: false,

“uncertain”: false,

“generic”: false,

“conditional”: false,

“historic”: false,

“temporality”: “”

},

{

“id”: “doc1_884540155_M_36”,

“classUri”: “Mass”,

“confidence”: 0.0,

“noteId”: “doc1_884540155”,

“begin”: 899,

“end”: 903,

“negated”: false,

“uncertain”: false,

“generic”: false,

“conditional”: false,

“historic”: false,

“temporality”: “”

},

{

“id”: “doc1_884540155_M_5”,

“classUri”: “Mass_In_Breast”,

“confidence”: 0.0,

“noteId”: “doc1_884540155”,

“begin”: 2007,

“end”: 2018,

“negated”: false,

“uncertain”: false,

“generic”: false,

“conditional”: false,

“historic”: false,

“temporality”: “”

},

{

“id”: “doc1_884540155_M_38”,

“classUri”: “Mass”,

“confidence”: 0.0,

“noteId”: “doc1_884540155”,

“begin”: 1107,

“end”: 1111,

“negated”: false,

“uncertain”: false,

“generic”: false,

“conditional”: false,

“historic”: false,

“temporality”: “”

},

{

“id”: “doc1_884540155_M_21”,

“classUri”: “Mass”,

“confidence”: 0.0,

“noteId”: “doc1_884540155”,

“begin”: 428,

“end”: 432,

“negated”: false,

“uncertain”: false,

“generic”: false,

“conditional”: false,

“historic”: false,

“temporality”: “”

},

{

“id”: “doc1_884540155_M_18”,

“classUri”: “Epithelial_Ovarian_Cancer”,

“confidence”: 0.0,

“noteId”: “doc1_884540155”,

“begin”: 1432,

“end”: 1449,

“negated”: false,

“uncertain”: false,

“generic”: false,

“conditional”: false,

“historic”: true,

“temporality”: “”

},

{

“id”: “doc1_884540155_M_2”,

“classUri”: “Mass”,

“confidence”: 0.0,

“noteId”: “doc1_884540155”,

“begin”: 1215,

“end”: 1219,

“negated”: false,

“uncertain”: false,

“generic”: false,

“conditional”: false,

“historic”: false,

“temporality”: “”

}

],

“indirectEvidence”: [],

“notEvidence”: [],

“confidenceFeatures”: []

},

{

“id”: “Mass_In_Breast_1683124727183”,

“classUri”: “Mass_In_Breast”,

“name”: “tumor_type”,

“value”: “PrimaryTumor”,

“directEvidence”: [

{

“id”: “doc1_884540155_M_22”,

“classUri”: “Mass”,

“confidence”: 0.0,

“noteId”: “doc1_884540155”,

“begin”: 528,

“end”: 532,

“negated”: false,

“uncertain”: false,

“generic”: false,

“conditional”: false,

“historic”: false,

“temporality”: “”

},

{

“id”: “doc1_884540155_M_3”,

“classUri”: “Mass_In_Breast”,

“confidence”: 0.0,

“noteId”: “doc1_884540155”,

“begin”: 402,

“end”: 415,

“negated”: false,

“uncertain”: true,

“generic”: false,

“conditional”: false,

“historic”: true,

“temporality”: “”

},

{

“id”: “doc1_884540155_M_23”,

“classUri”: “Lobular_Mass”,

“confidence”: 0.0,

“noteId”: “doc1_884540155”,

“begin”: 751,

“end”: 765,

“negated”: false,

“uncertain”: false,

“generic”: false,

“conditional”: false,

“historic”: false,

“temporality”: “”

},

{

“id”: “doc1_884540155_M_20”,

“classUri”: “Dysplastic_Nevus”,

“confidence”: 0.0,

“noteId”: “doc1_884540155”,

“begin”: 1650,

“end”: 1653,

“negated”: false,

“uncertain”: false,

“generic”: false,

“conditional”: false,

“historic”: false,

“temporality”: “”

},

{

“id”: “doc1_884540155_M_35”,

“classUri”: “Mass_In_Breast”,

“confidence”: 0.0,

“noteId”: “doc1_884540155”,

“begin”: 2039,

“end”: 2052,

“negated”: false,

“uncertain”: false,

“generic”: false,

“conditional”: false,

“historic”: false,

“temporality”: “”

},

{

“id”: “doc1_884540155_M_36”,

“classUri”: “Mass”,

“confidence”: 0.0,

“noteId”: “doc1_884540155”,

“begin”: 899,

“end”: 903,

“negated”: false,

“uncertain”: false,

“generic”: false,

“conditional”: false,

“historic”: false,

“temporality”: “”

},

{

“id”: “doc1_884540155_M_5”,

“classUri”: “Mass_In_Breast”,

“confidence”: 0.0,

“noteId”: “doc1_884540155”,

“begin”: 2007,

“end”: 2018,

“negated”: false,

“uncertain”: false,

“generic”: false,

“conditional”: false,

“historic”: false,

“temporality”: “”

},

{

“id”: “doc1_884540155_M_38”,

“classUri”: “Mass”,

“confidence”: 0.0,

“noteId”: “doc1_884540155”,

“begin”: 1107,

“end”: 1111,

“negated”: false,

“uncertain”: false,

“generic”: false,

“conditional”: false,

“historic”: false,

“temporality”: “”

},

{

“id”: “doc1_884540155_M_21”,

“classUri”: “Mass”,

“confidence”: 0.0,

“noteId”: “doc1_884540155”,

“begin”: 428,

“end”: 432,

“negated”: false,

“uncertain”: false,

“generic”: false,

“conditional”: false,

“historic”: false,

“temporality”: “”

},

{

“id”: “doc1_884540155_M_18”,

“classUri”: “Epithelial_Ovarian_Cancer”,

“confidence”: 0.0,

“noteId”: “doc1_884540155”,

“begin”: 1432,

“end”: 1449,

“negated”: false,

“uncertain”: false,

“generic”: false,

“conditional”: false,

“historic”: true,

“temporality”: “”

},

{

“id”: “doc1_884540155_M_2”,

“classUri”: “Mass”,

“confidence”: 0.0,

“noteId”: “doc1_884540155”,

“begin”: 1215,

“end”: 1219,

“negated”: false,

“uncertain”: false,

“generic”: false,

“conditional”: false,

“historic”: false,

“temporality”: “”

}

],

“indirectEvidence”: [],

“notEvidence”: [],

“confidenceFeatures”: []

},

{

“id”: “historic_1683124727183”,

“classUri”: “historic”,

“name”: “historic”,

“value”: “historic”,

“directEvidence”: [],

“indirectEvidence”: [],

“notEvidence”: [],

“confidenceFeatures”: [],

“confidence”: 10

},

{

“id”: “calcifications_1683124727183”,

“classUri”: “calcifications”,

“name”: “calcifications”,

“value”: “false”,

“directEvidence”: [],

“indirectEvidence”: [],

“notEvidence”: [],

“confidenceFeatures”: [],

“confidence”: 10

},

{

“id”: “_1683124727183”,

“classUri”: “”,

“name”: “grade”,

“value”: “9”,

“directEvidence”: [],

“indirectEvidence”: [],

“notEvidence”: [],

“confidenceFeatures”: []

}

]

}

]

}

